# Altered Intercellular Communication and Extracellular Matrix Signaling as a Potential Disease Mechanism in Human Hypertrophic Cardiomyopathy

**DOI:** 10.1101/2021.12.18.21268004

**Authors:** Amy Larson, Christina J. Codden, Gordon S. Huggins, Hassan Rastegar, Frederick Y. Chen, Barry J. Maron, Ethan J. Rowin, Martin S. Maron, Michael T. Chin

## Abstract

**Objectives:** To understand Hypertrophic Cardiomyopathy-associated alterations in gene expression and intercellular communication at the single cell level in left ventricular outflow tract lesions.

**Background:** Human hypertrophic cardiomyopathy (HCM) is considered a disorder of the sarcomere (i.e., cardiomyocytes) but the paradoxical association of nonmyocyte phenotypes such as fibrosis, mitral valve anomalies and microvascular occlusion is unexplained.

**Methods:** To understand the interplay between cardiomyocyte and nonmyocyte cell types in human HCM, single nuclei RNA-sequencing (snRNA-seq) was performed on myectomy specimens from HCM patients with left ventricular outflow tract obstruction and control samples from donor hearts free of cardiovascular disease.

**Results:** Clustering analysis identified a total of 34 distinct cell populations, which were classified into 10 different cell types based on marker gene expression. Differential gene expression analysis comparing HCM to Normal datasets revealed differences in sarcomere and extracellular matrix gene expression. Analysis of expressed ligand-receptor pairs across multiple cell types indicated profound disruption in HCM intercellular communication, particularly between cardiomyocytes and fibroblasts, fibroblasts and lymphocytes and involving integrin β1 and its multiple extracellular matrix (ECM) cognate ligands.

**Conclusions:** These findings provide evidence for intercellular interactions in HCM that link sarcomere dysfunction with altered cardiomyocyte secretion of ECM ligands, altered fibroblast ligand-receptor interactions with a variety of cell types and increased fibroblast to lymphocyte signaling, which can further alter the ECM composition, disrupt cellular function and promote nonmyocyte phenotypes.

## Introduction

Hypertrophic cardiomyopathy (HCM) is an autosomal dominant inherited disorder characterized by unexplained left ventricular hypertrophy, often asymmetric in nature. HCM is frequently complicated by diastolic heart failure, left ventricular outflow tract (LVOT) obstruction, ventricular tachyarrhythmias, sudden cardiac death, microvascular angina, and atrial fibrillation (reviewed in ^1^). Approximately 70% of patients who present clinically with HCM have some degree of LVOT tract obstruction, commonly from asymmetric hypertrophy of the interventricular septum (IVS). LVOT obstruction is the most concordant phenotype aside from unexplained left ventricular hypertrophy. Histologically, myocyte disarray, cardiac fibrosis, and microvascular occlusion are found. Genetic studies have identified numerous mutations in a variety of sarcomere genes such as *MYH7, MYL2, MYL3, MYBPC3, TNNT2, TNNI3* and *TPM1*, leading to the concept that HCM is a disease of the sarcomere ^2^. Among patients with sarcomere gene mutations, the most commonly affected gene is *MYBPC3*, followed closely by *MYH7*. Together *MYBPC3* and *MYH7* mutations account for approximately 70% of patients with sarcomere mutations. Genetic studies have also identified additional mutations in non-sarcomere genes. Such mutations can indirectly lead to sarcomere dysfunction and consequently HCM ^3, 4^.

Despite significant advances in understanding the genetics of HCM, genetic testing is only informative in approximately 30% of probands, and it has been recently reported that polygenic factors contribute to the phenotype ^5-7^. Among patients and families, there is often variable penetrance, resulting in a wide spectrum of disease even in families with the same mutation, suggesting the influence of modifier genes and environmental factors. At the molecular and cellular level, downstream pathway alterations, secondary to sarcomere dysfunction, that predispose to HCM complications (e.g., LVOT obstruction, sudden cardiac death, cardiac fibrosis, microvascular dysfunction) are poorly understood especially in the context of intact mature cells within hypertrophic, obstructive lesions. We hypothesize that there are final common, cell-specific pathological pathways independent of specific genetic origin that promote the concordant LVOT obstruction phenotype in HCM patients. A recent proteomic study supports convergent proteomic mechanisms ^8^. We also hypothesize that activation of pathological pathways in noncardiomyocytes, presumably in response to effects on cardiomyocytes, is relevant to the HCM phenotype. To date, no studies of this nature have been conducted in the HCM population and no common final pathological pathways driving LVOT obstruction have been identified. Identification of novel, cell type-specific pathological alterations in gene expression will provide insight into the transcriptional programs activated in LVOT obstructive lesions and may link HCM to other mechanisms of pathological hypertrophy. Determination of pathological pathway activation will also facilitate the identification of potential targets for future novel therapies for HCM.

Advances in single cell RNA-sequencing have allowed examination of single cell transcription patterns in tissues, thereby allowing the identification of cell subpopulations that contribute to cancer and tissue renewal. Single cell transcriptomics have also provided insight into intracellular molecular biology and interactions between cell populations in specific tissues. Single cell analyses of heart tissue in mouse models have revealed important insights into cardiovascular development, identification of distinct cardiomyocyte subpopulations ^9-11^, and adult cardiovascular disease ^12^. Single cell analyses of normal human cardiac tissue have documented the regional cellular and transcriptional diversity of the human heart ^13-15^ but only two have assessed the IVS ^14, 15^. We have performed a single nuclei analysis of gene expression in heart IVS tissue from a cohort of HCM patients of varying genotypes and unused donor heart controls to identify candidate common pathway regulators of the HCM phenotype. We have identified cell type-specific alterations in gene expression, with changes in contractile protein gene expression in cardiomyocytes and in extracellular matrix (ECM) gene expression in cardiac fibroblasts. Analysis of ligand-receptor (L-R) pair gene expression, which outlines the potential for intercellular communication, reveals a striking decrease in ligands and receptors associated with HCM and integrin-β1 (ITGβ1) signaling, thus providing evidence linking sarcomere dysfunction with extracellular matrix signaling and thereby suggesting a mechanistic link between cardiomyocyte function and noncardiomyocyte manifestations of HCM.

## Methods

### Study Patients, Sample Collection and Processing

A total of 9 patients with clinically documented HCM scheduled for surgical myectomy were approached for written informed consent to allow their tissue to be used for research. Those who consented underwent myectomy and tissue was collected. Myectomy sample processing was done as previously described ^15^. 100 mg of collected myectomy tissue was minced into 1 mm^3^ pieces, placed in 0.5 mL of CryoStor CS10 Freeze Media (STEMCELL Technologies), and stored in a MrFrosty (ThermoFisher) at 4°C for 10 minutes and then transferred to −80°C overnight. Bulk RNA was isolated from a piece of tissue using the Qiagen RNeasy Plus Micro kit and then assessed on the Agilent Bioanalyzer 2100. Samples with an RNA Integrity Number greater than 8.5 were used in library preparation. Sample collection was approved by the Tufts University/Medical Center Health Sciences Institutional Review Board under IRB protocol # 9487. All subjects gave their informed consent for inclusion before they participated in the study. The study was conducted in accordance with the Declaration of Helsinki. Patient characteristics were obtained from the medical record and are shown in Supplemental Table ST1. Tissue from organ donor patients without underlying cardiac disease was obtained and processed as described previously ^15^.

### Nuclei isolation, library preparation, and sequencing

Generation of single nuclei sequencing libraries was performed as previously described ^15^. Cryopreserved samples were thawed at 37°C and placed on ice. Nuclei were isolated via Dounce homogenization as previously described ^16^. Homogenates were filtered through a Pluristrainer 10 µM cell strainer (Fisher Scientific) into a pre-chilled tube. Nuclei were pelleted by centrifuging at 500 x g for 5 min at 4°C. Nuclei pellets were washed and pelleted according to manufacturer protocol (10x Genomics). Nuclei were stained with trypan blue and counted on a hemocytometer to determine concentration prior to loading of the 10x Chromium device and samples were diluted to capture ∼10,000 nuclei. Nuclei were separated into Gel Bead Emulsion droplets using the 10x Chromium device according to the manufacturer protocol (10x Genomics). Sequencing libraries were prepared using the Chromium Single Cell 3’ reagent V2 kit according to manufacturer’s protocol. Libraries were multiplexed and sequenced on a NextSeq550, NovaSeq S2, or NovaSeq S4 (Illumina) to produce ∼50,000 reads per nucleus. Single nuclei RNAseq data for normal IVS tissue is available in the Gene Expression Omnibus database under accession number GSE161921 ^15^. Data for HCM IVS tissue is available under accession number GSE174691.

### Clustering of Cells by Gene Expression Pattern and Assignment of Cell Type Identity

Sequencing reads were processed using Cell Ranger 6.0.1 ^17^. The gene expression matrix was subset to only include reads from the nuclear genome. Quality control (QC) filtering, clustering, dimensionality reduction, visualization, and differential gene expression were performed using the R package Seurat 3.5.0. Each dataset was filtered so that genes that were expressed in three nuclei or more were included in the final dataset. The dataset was further sublet to exclude nuclei that had fewer than 200 genes expressed to remove droplets containing only ambient RNA, and to exclude nuclei with greater than 2000 genes to remove droplets that contained two nuclei. Datasets were individually normalized and integrated using Seurat’s SCTransform development workflow to reduce batch effects ^18^. Optimal clustering resolution was determined using Clustree ^19^ to identify the resolution where the number of clusters stays stable and was determined to be 0.9 for the integrated dataset. Expression of known cell-specific gene markers were used to identify major cell types. Differentially expressed genes and their functions were used to identify clusters without an assigned identity, or to further refine the cell type.

### Trajectory Analysis and Identification of Differentially Expressed Genes

Trajectory analysis was performed using Monocle3 ^20^ to determine the relationship between subtypes of cells identified in our clustering analysis. Since our data do not represent a developmental time course, we determined the root nodes for each subtype by hierarchical clustering prior to generating trajectories and assigning pseudotime to each cell. Each cell type was analyzed in three cohorts: 1. Normal and HCM cells together; 2. Normal cells alone; 3. HCM cells alone. Differentially expressed genes over trajectory paths in uniform manifold approximation and projection (UMAP) space (i.e., spatial autocorrelation) was performed in Monocle3 using Moran’s I statistic. Moran’s I statistic is a value that varies from −1 to 1, where - 1 indicates perfect dispersion, 0 indicates no spatial autocorrelation, and 1 indicates perfect positive autocorrelation (i.e., nearby cells in have similar gene expression values in focal region of UMAP space). For each Normal and HCM cell type, a gene was determined to be differentially expressed over space if the associated Moran’s I statistic value was positive, paired with a significant adjusted p-value ≤ 0.05, and expressed in ≥ 1% of associated cells. Since many genes showed differential expression over space, further conservative filtering was performed in which genes with Moran’s I statistic available in a single class (i.e., Normal or HCM) were filtered by Moran’s I statistic values >0.1. For genes with Moran’s I statistics available in both classes (i.e., Normal and HCM), genes were filtered by an absolute difference >0.1. Gene Ontology (GO) analysis of molecular function and biological process associated with differentially expressed genes was done using the online tools at uniprot.org/ uniprotkb ^21^.

### Analysis of Ligand-Receptor Pair Gene Expression to Discover Intercellular Communication Pathways

To quantify potential cardiac cell-cell communication in Normal and HCM hearts, cell communication networks were plotted in igraph ^22^ and compared on the basis of ligand-receptor pair gene expression. Our cell-cell communication networks were derived as described previously ^11^, using a list of 2557 human ligand-receptor pairs ^23^. A ligand or receptor for each cell type or cluster was considered expressed if the corresponding gene showed an above zero gene count in ≥ 20% of cells in our snRNAseq data. We initially analyzed the potential signaling interactions between the 10 cell types identified in our snRNAseq data. Lines in our cell networks connect two cell types and represent expressed human ligand-receptor pairs (i.e., potential cell-cell communication between a broadcasting (ligand) and recipient (receptor) cell types. Line color in our networks represents the broadcasting ligand source. Line thickness is proportional to the number of uniquely expressed ligand-receptor pairs. Cell-cell communication networks were also analyzed by fibroblast cluster along with other cell types and also by fibroblast clusters and cardiomyocyte clusters. GO analysis of differentially expressed ligand receptor pairs was performed using the R package clusterProfiler ^24^.

### Statistics

#### Power calculations

We used mixed effects models to analyze cell type specific differential expression, while taking into account the variability between and within subjects. A sample size of 6 cases and 6 controls, with an average of 3,000 cells per subject, will provide 80% power to detect fold change of expression ranging between 1.3 for an intracluster correlation f 0.01, and 2 for an intracluster correlation of 0.1. The power calculations were pretty stable for sample sizes ranging between 500 and 3000 cells per subject. We used a Bonferroni correction for 10,000 tests to fix the level of significance. The power calculations were conducted using Power Analysis and Sample Size software (PASS).

Other statistical methods to cluster cells in UMAP space and control for batch effects using the built-in functionality of Seurat ^18^, to determine cluster stability by Clustree ^19^ and to perform Gene Ontology Analysis using well documented software programs ^24^ are described in above methods sections and in cited references. Statistical methods to compare gene expression along pseudotime trajectories using linear regression or spatial autocorrelation using the built-in functionality of Monocle3 are described above and in the original cited reference ^20^.

## Results

### Patient characteristics

Surgical myectomy tissue was obtained from nine HCM patients with severely symptomatic LVOT obstruction. Patient characteristics are summarized in Supplemental Table ST1. Patients gave informed consent for their myectomy tissue to be used in research. The patients varied in age from 37 to 76. Three of nine were female. One of the patients carried a pathogenic Mybpc3 mutation, 1 patient had a likely pathogenic KRAS mutation, and the remaining seven patients had no known mutations pathogenic for HCM. The patients and datasets for the normal human IVS have been previously described ^15^.

### Single nuclei RNA-sequencing of human HCM myectomy tissue reveals extensive cardiomyocyte and fibroblast diversity in the interventricular septum but does not reveal a disease-specific population of cells

After sequencing and initial data processing with Cell Ranger software ^17^, each sample dataset was processed further to remove called nuclei that were likely droplets with only ambient RNA, or droplets that contained two nuclei. The nine HCM datasets and four donor heart datasets were combined into one dataset using the Seurat Integration function ^18^. The final combined dataset included 109,823 nuclei from HCM hearts and 24,858 nuclei from donor hearts. Clustering of the integrated dataset revealed 34 cell populations within the IVS, and this was visualized using the dimensionality reduction algorithm UMAP (Fig. 1A), where each point represents a single nucleus colored by cluster identity. Visualizing the integrated dataset by Normal and HCM datasets reveals that all 34 clusters are present in each case, with no cell populations (i.e., clusters) specific to either condition (Fig. 1B).

**Figure 1.**
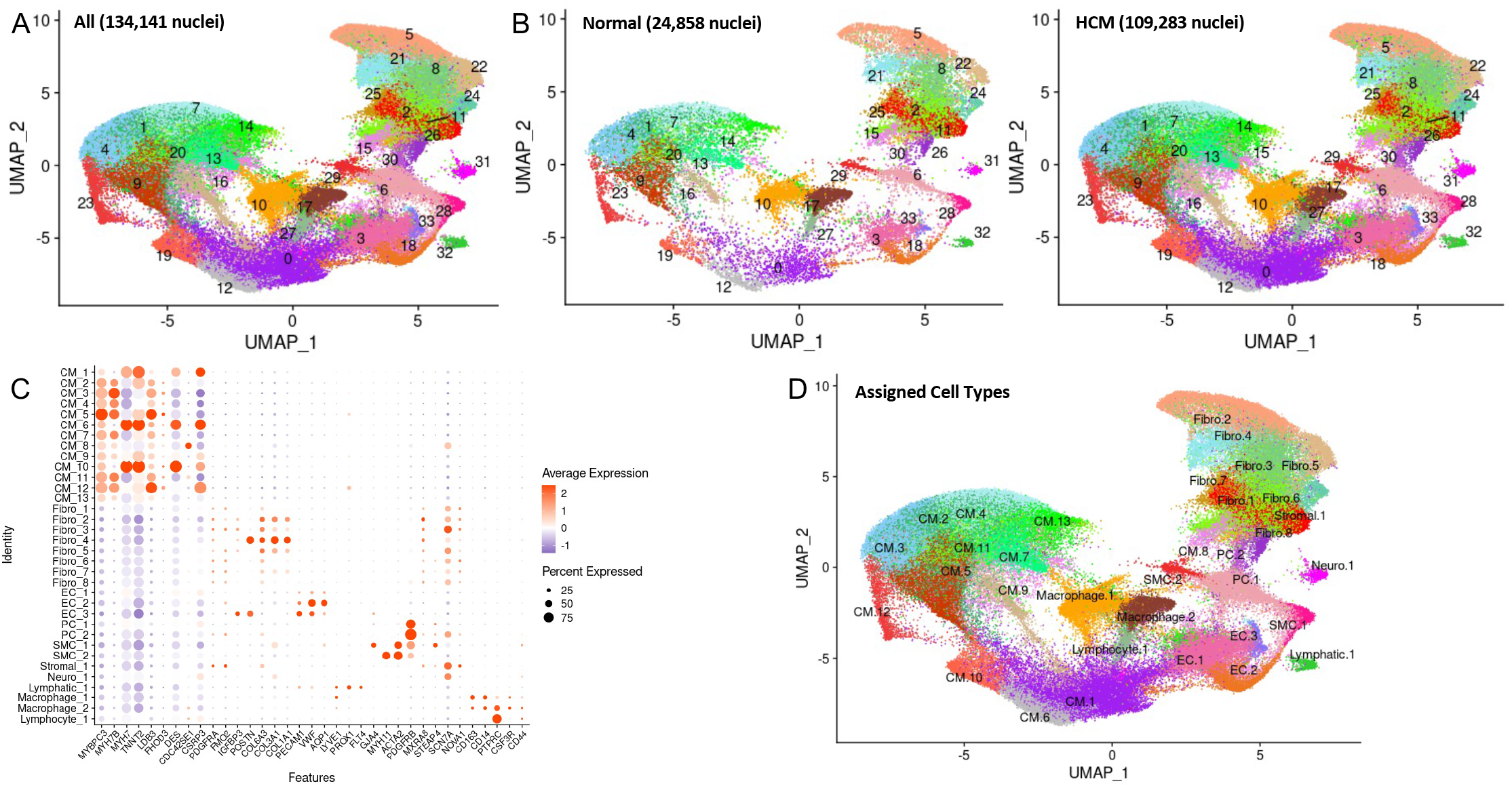
Single nuclei RNA-seq of Normal and HCM IVS Tissue Reveals Cellular Diversity but No HCM-associated Cell Types. **A**. UMAP plot of 34 distinct cell clusters identified in the combined Normal and HCM dataset. **B**. Separating the cell clusters by disease label does not identify HCM-specific cell types. **C**. Listing of cell markers used to assign cell types. **D**. UMAP plot of combined dataset showing cell identity labels instead of cluster numbers.

Cell identities were assigned to each cluster using known gene markers of expected cell types, differentially expressed genes, gene ontology using GOstats ^25^, and Ingenuity Pathway Analysis ^26^. Figure 1C lists the gene markers used to identify each cell type and illustrates each cluster’s unique expression of gene markers. Upon assigning cell types, similar cell types were expectedly positioned close to each other in UMAP space (Fig. 1D). Interestingly, we see thirteen separate cardiomyocyte populations and eight different fibroblast populations, revealing significant cardiomyocyte and fibroblast diversity in the IVS (Fig. 1C, 1D). Other cell types identified included endothelial, lymphatic, smooth muscle, pericyte, neuronal, stromal, and immune cell populations (macrophages, lymphocytes). Among normal donor heart samples, assigned cardiomyocytes made up approximately 41.0% (mean, range 30.1 to 61.6%) of the total cell population. Similarly, among HCM myectomy samples cardiomyocytes made up approximately 51.6% (mean, range 35.5-63.0%, p-value not significant) of the total cell population. Noncardiomyocytes make up the remaining cell population. HCM does not appear to be associated with a shift in the relative proportion of cells in the heart.

### Trajectory and differential gene expression analysis reveals HCM-associated perturbations

Clustering analysis revealed great diversity for both cardiomyocytes and fibroblasts. To gain insight into potential relationships among the different cell populations for each cell type, clusters were projected onto pseudotime trajectory analysis using Monocle3 ^20^. Trajectory analysis was performed on Normal cells only, HCM cells only, and both conditions combined for each of the 10 cell types identified in the above analysis. In single-cell trajectory analysis, a trajectory is a computed path that describes a cell type’s biological progression through a dynamic process. Prior to building trajectory paths, the root node, or beginning, of each trajectory path was determined through hierarchical clustering of our single nuclei data. Then, trajectory paths for each assigned cell type (assigned in Seurat) were constructed in UMAP space using Monocle3 with Normal and HCM data together and individually. Once trajectory paths were established, pseudotime could be assigned to each cell. For all cell types, root nodes showed consistent placement among Normal only groups, HCM only groups, and Normal and HCM groups. Trajectory paths also showed similar basic structures among Normal only groups, HCM only groups, and Normal and HCM groups. Small differences in trajectory paths for a single cell type among groups are likely due to different sample sizes in Normal and HCM groups (e.g., Normal cardiomyocyte nuclei n=10131, HCM cardiomyocyte nuclei n=57847). Therefore, we were not able to distinguish any meaningful changes among trajectory paths. Representative trajectories and root nodes are shown for the two most common cell types, cardiomyocytes and fibroblasts, in Supplemental Figure S1. These findings suggest that the relationships between the various subtypes of each cell type do not vary significantly in HCM heart tissue in comparison to Normal heart tissue.

Establishment of trajectories facilitates the analysis of differential gene expression along the trajectory and between conditions along the trajectory. We analyzed differential gene expression for each cell type along their trajectories for the Normal and HCM populations through spatial autocorrelation. Moran’s I statistic and adjusted p-values were calculated for each gene in Normal only and HCM only groups within each cell type. When paired with a significant p-value (≤0.05), a Moran’s I statistic value near zero indicated no spatial autocorrelation and a value near 1 indicated perfect positive autocorrelation in which a gene is expressed in a focal region of the UMAP space. Results showed between 91 and 3589 genes are differentially expressed along trajectory paths among cell types and between 4 and 1823 differentially expressed genes overlapped between Normal and HCM groups among cell types (Supplemental Table ST2).

Since many genes showed differential expression over space, further conservative filtering was performed to identify genes of interest. Genes with Moran’s I statistic available for a single condition, Normal or HCM, were filtered by a Moran’s I statistic value above 0.1, while genes with Moran’s I statistics available for both classes were filtered by an absolute difference >0.1. For each cell type, 0 to 41 genes passed the filter with 176 unique genes in total passing the filter among all cell types (Supplemental Table ST3). The expression of these filtered genes was then plotted in UMAP space for their associated cell type in Normal and HCM conditions as shown for a subset of identified genes in cardiomyocytes (Supplemental Figure S2). Visual analysis of the UMAP plots revealed 28 genes with pronounced differences in their spatial expression between Normal and HCM conditions (summary in Table 1). Of these 28 genes, 6 (*ACTA1, ACTC1, MYHL7, MYL2, TNNI3, TNNI2*) are already known to be associated with human HCM. Extracellular matrix genes (*COL1A1, COL6A1, COL6A2, CYR61, POSTN*) are also differentially expressed in Normal vs HCM fibroblasts, suggesting a change in ECM composition. GO Enrichment analysis of these 28 differentially expressed genes revealed significant enrichment for molecular functions involving the ECM, biological processes involving muscle development and cellular components involving the ECM (Supplemental Fig. S3A). Analysis of genes from this list showing increased expression in obstructive HCM revealed significant enrichment in molecular functions involving actin binding, biological processes involving muscle filament sliding, contraction and movement and cellular components involving the sarcomere, myofibril and contractile fiber (Supplemental Fig. S3B). Analysis of genes from this list showing decreased expression in HCM revealed enrichment for molecular functions involving the ECM, biological processes involving the response to transforming growth factor-β (TGF-β), steroid hormones, the immune response and ECM organization, and cellular components involving the ECM (Supplemental Fig. S3C).

**Table 1.** Differentially Expressed Genes in Normal and HCM IVS Cells.

|  | **Gene of Interest** | **Affected Cell Group** | **Expression in HCM** | **Established HCM Gene** |
| --- | --- | --- | --- | --- |
| 1 | ACTA1 | Neuronal | Increased |  |
| 2 | ACTC1 | Cardiomyocyte | Increased | Yes |
| 3 | ADH1B | Cardiomyocyte cluster 8 | Decreased |  |
| 4 | C1R | Fibroblast | Decreased |  |
| 5 | C1S | Fibroblast | Decreased |  |
| 6 | C7 | Fibroblast | Decreased |  |
| 7 | CCL2 | Neuronal | Decreased |  |
| 8 | CFD | Endothelial cluster 1 | Increased |  |
| 9 | CILP | Fibroblast | Decreased |  |
| 10 | COL1A1 | Fibroblast | Decreased |  |
| 11 | COL6A1 | Fibroblast | Decreased |  |
| 12 | COL6A2 | Fibroblast, Lymphatic | Decreased |  |
| 13 | CYR61 | Fibroblast clusters 2, 4, 7 | Decreased |  |
| 14 | FOS | Leukocyte | Decreased |  |
| 15 | HES1 | Fibroblast | Decreased |  |
| 16 | IGFBP7 | Fibroblast | Decreased |  |
| 17 | JUNB | Cardiomyocyte | Decreased |  |
| 18 | MFAP4 | Fibroblast | Decreased |  |
| 19 | MS4A6A | Cardiomyocyte | Decreased |  |
| 20 | MYH7 | Cardiomyocyte | Increased | Yes |
| 21 | MYL2 | Myofibroblast, Smooth Muscle, Neuronal | Increased | Yes |
| 22 | NEAT1 | Cardiomyocyte, Cardiomyocyte cluster 1 & 6, Pericyte | Decreased |  |
| 23 | PMEPA1 | Neuronal | Decreased |  |
| 24 | POSTN | Fibroblast, Endothelial | Decreased |  |
| 25 | S100A1 | Smooth Muscle | Increased |  |
| 26 | SERPINF1 | Fibroblast | Decreased |  |
| 27 | TNNI3 | Cardiomyocyte, Smooth Muscle | Increased | Yes |
| 28 | TNNT2 | Myofibroblast, Lymphatic | Increased | Yes |

### Analysis of ligand-receptor pair gene expression indicates profound alteration of intercellular communication in HCM

#### HCM is associated with a general decrease in cell-cell communication but an increase in fibroblast to lymphocyte communication

To quantify potential cardiac cell-cell communication in Normal and HCM IVS tissue, we quantified the number of possible expressed ligand-receptor pairs among cell types as previously described ^11^. We examined the expression of a curated list of 2557 human ligand-receptor pairs ^23^ in which ligands and receptors were considered expressed if their associated gene was detectable in ≥ 20% of cells in a cell type. Notably, quantification of cell-cell communication in this study represents potential communication as we account for only expressed ligand-receptor pairs and not the position or boundaries of cell types. Results showed that Normal cardiac IVS tissue demonstrated an overall dense intercellular communication network among our 10 identified cell types (Fig 2). HCM cardiac tissue, however, unexpectedly demonstrated an overall sparse intercellular communication network in which potential communication was drastically reduced compared to the Normal tissue (Fig 2A). Quantitatively, the total number of expressed ligand-receptor pairs in the HCM tissue was more than two times less than in Normal tissue (546 and 1138 respectively, Fig. 2B). Analysis of the broadcast ligands by individual cell types demonstrates that 1) cardiomyocytes show a broad decrease in both paracrine and autocrine ligand broadcasting in HCM (Fig. 2C, D) and 2) fibroblasts broadcast the greatest number of ligands to the other cell types in both Normal and HCM tissue. Fibroblast communication is particularly high with fibroblast, endothelial, pericyte, smooth muscle, neuronal, and cardiomyocyte cells (Fig. 2). While intercellular communication was generally reduced in HCM tissue compared to Normal tissue, there was one case of increased HCM communication from fibroblasts to lymphocytes (1 expressed ligand-receptor pairs in Normal tissue to 13 in HCM tissue; Fig 2C, D).

**Figure 2.**
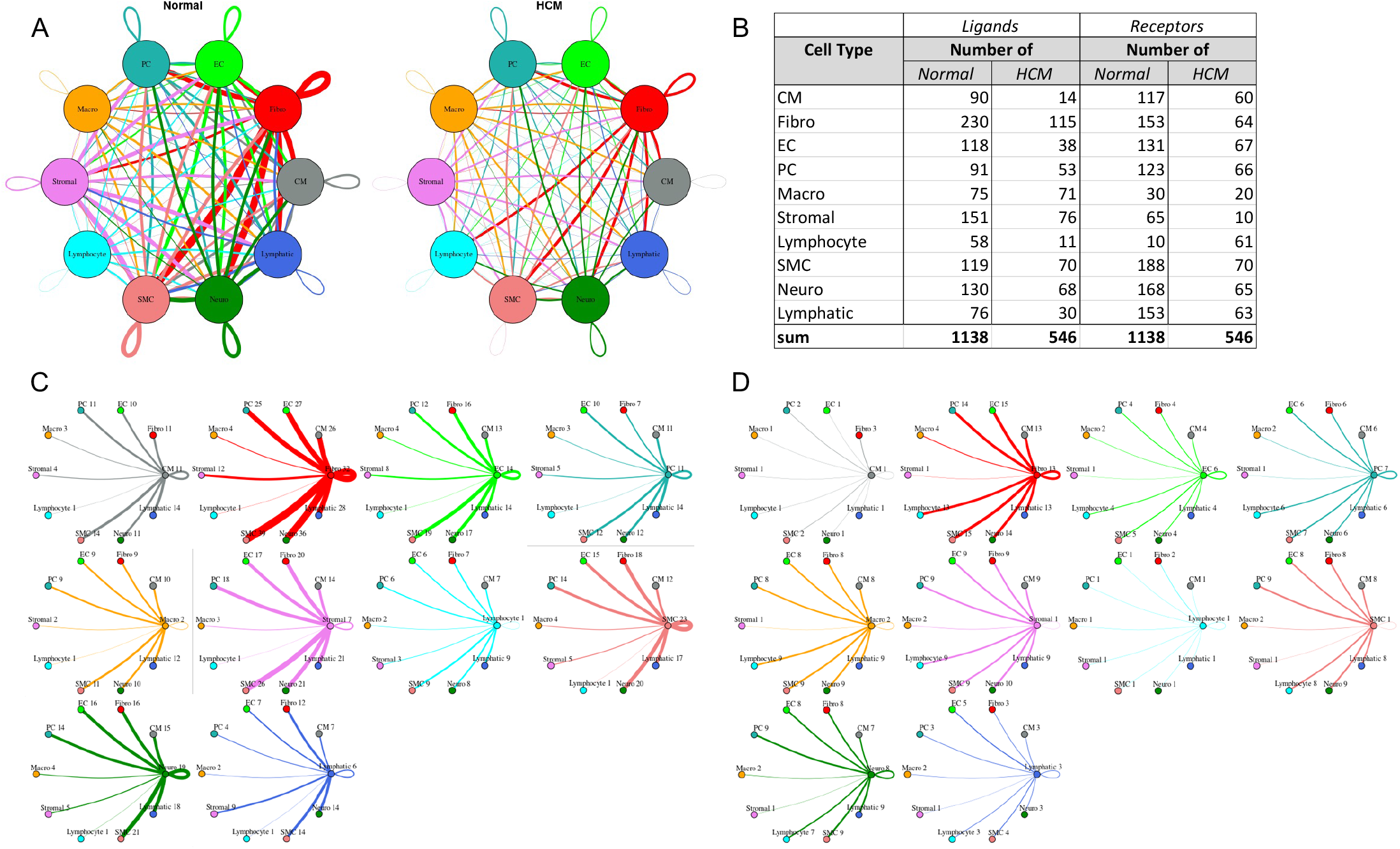
Intercellular communication networks are reduced in HCM. **A**. Cell-cell communication networks between cardiac cell types in normal control (left) and HCM (right) conditions. Line color indicates ligand broadcast by the cell population with the same color. Lines connect to cell types which expressed cognate receptors. Line thickness is proportional to the number of uniquely expressed ligand-receptor pairs. Loops indicates communication within a cell type. **B**. Quantity of ligands and receptors in expressed ligand-receptor pairs described by cell type and condition (Normal or HCM). **C**., **D**. Cell-cell communication networks broken down by cell type in normal control (**C**) and HCM (**D**) conditions. Figure formatting follows panel **A**. Numbers indicate the quantity of uniquely expressed ligand-receptor pairs between the broadcasting cell type (expressing ligand) and receiving cell type (expressing receptor).

To assess the molecular functions potentially affected by changes in ligand-receptor signaling among our 10 identified cell types, GO enrichment analysis was performed on the ligands and receptors in expressed ligand-receptor pairs from both Normal and HCM tissue (Fig. 3). We observed a global decrease in nearly all identified ligand functions in the HCM heart tissue compared to the Normal heart tissue, particularly in functions such as extracellular matrix structural constituent (55% total ligand count decrease), extracellular matrix structural constituent conferring tensile strength (54% total ligand decrease), growth factor binding (63% total ligand count decrease), protease binding (46% total ligand count decrease), and platelet-derived growth factor binding (63% total ligand count decrease; Fig. 3A). As expected, the molecular functions associated with changes in receptor expression mirrored those observed for changes in ligand expression (Supplemental Fig. S4A). HCM cell types including cardiomyocytes, endothelial, macrophages and lymphocytes showed disproportionately large decreases in the above listed ligand processes ranging from 67-100% decreases in total ligand counts associated with a process compared to Normal ligand counts (Figure 3B-E). Two of these ligand processes (growth factor binding and platelet-derived growth factor binding) were completely lost in the HCM condition in cardiomyocytes, endothelial, macrophages and lymphocytes (Figure 3B-E). These trends found in ligand GO enrichment analysis were also identified in receptor GO enrichment analysis, except in lymphocytes (Supplemental Fig. S4B-F). In Normal tissue, lymphocyte receptors do not overrepresent any GO molecular functions, but in HCM tissue they show overrepresentation of receptors associated with coreceptor activity, cytokine binding, exogenous protein binding, integrin binding, etc., suggesting important changes in lymphocyte function. Together, ligand and receptor findings suggest that growth, the extracellular matrix, protein breakdown and lymphocyte function may be highly affected in HCM cardiac cells. Since fibroblast to lymphocyte signaling was the only pathway associated with increased ligand-receptor interactions in HCM heart tissue, we examined the specific ligand-receptor interactions in fibroblast to lymphocyte signaling that were increased (Supplemental Table ST4). In HCM, lymphocytes gained *ITGB1* receptor expression, allowing communication with several cognate ligands expressed in fibroblasts (*COL1A1, COL1A2, COL3A1, COL6A1, COL6A2, COL6A3, FBLN1, FN1, HSPG2, VCAN*). HCM fibroblasts also gained expression of *COL1A1* and *COL1A2* ligands which enabled communication with the *CD36* lymphocyte receptor. These findings further underscore the importance of the extracellular matrix in HCM.

**Figure 3.**
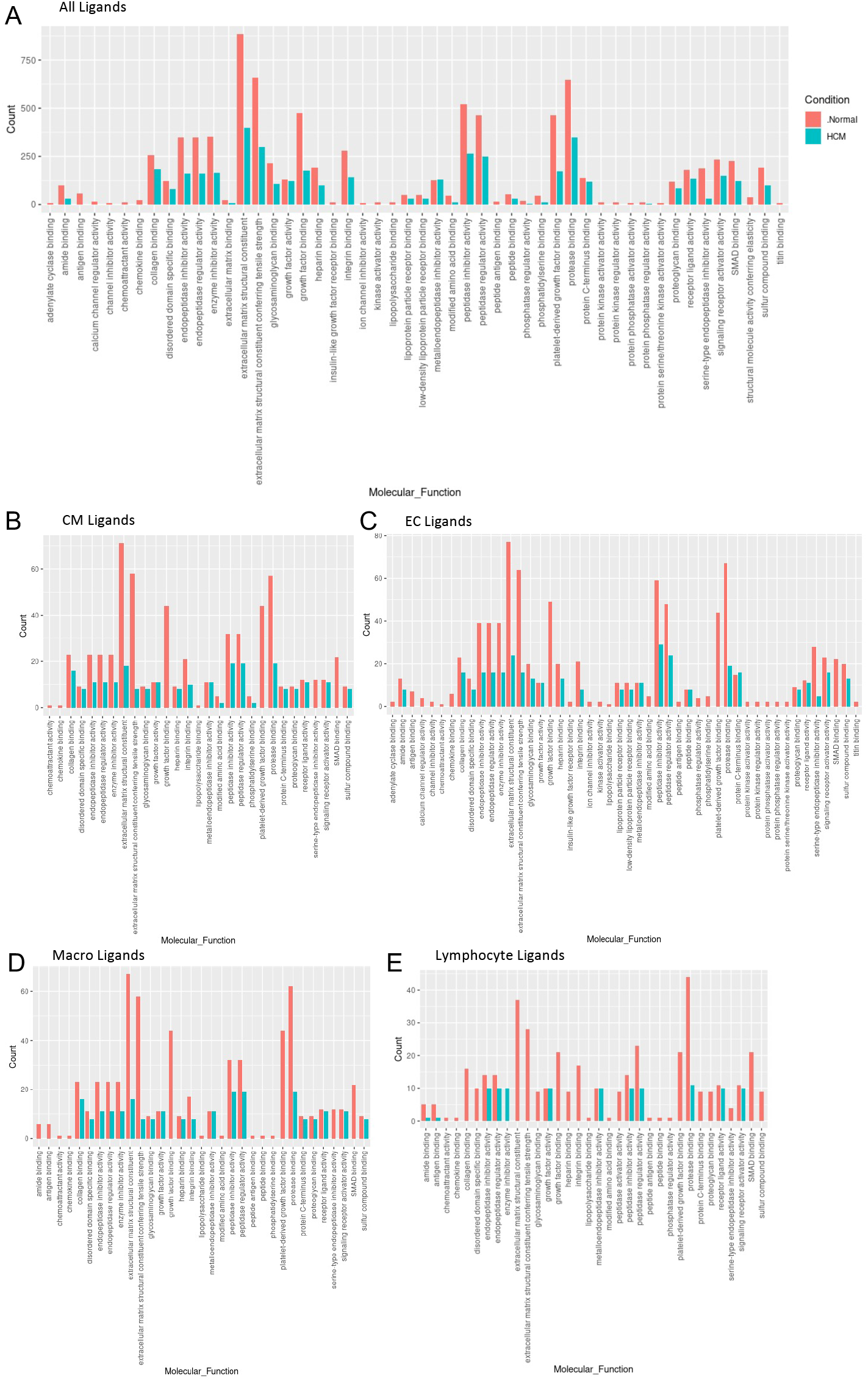
Bar plot representing the total count of ligands (in expressed ligand-receptor pairs) associated with different cellular processes in Normal and HCM IVS Cells. Bar color distinguishes ligand count in normal or HCM conditions. **A**. Comparison of molecular processes across all cell types. **B**. Comparison in cardiomyocytes. **C**. Endothelial cells. **D**. Macrophages. **E**. Lymphocytes.

#### Fibroblast subtypes show subtype-specific changes in cell-cell communication

To better understand the interaction between the different fibroblast populations and other cell types in the IVS, we examined the potential ligand-receptor communication through cell network plots for all fibroblast subtypes alongside the other previously identified 9 cell types (Fig. 4A, C, D). In Normal tissue, all fibroblast subtypes broadcast ligands extensively. In HCM tissue, fibroblast subtypes show a drastically reduced number of broadcasting ligands compared to Normal tissue, (1694 versus 4573 respectively; Fig. 4). There was a disproportionate reduction in *ITGB1* and *LRP1* receptor expression in fibroblast clusters 7 and 8 (Fig. 4B), which disabled communication with several ligands (ITGβ1 to COL1A1, COL1A2, COL3A1, COL6A1, COL6A3, FBLN1, FN1, HSPG2, and VCAN ligands; LRP1 to APP, C3, CALR, CTGF, HSP90B1, LRPAP1, PSAP, SERPING1, SERPINE2, TFPI).

**Figure 4.**
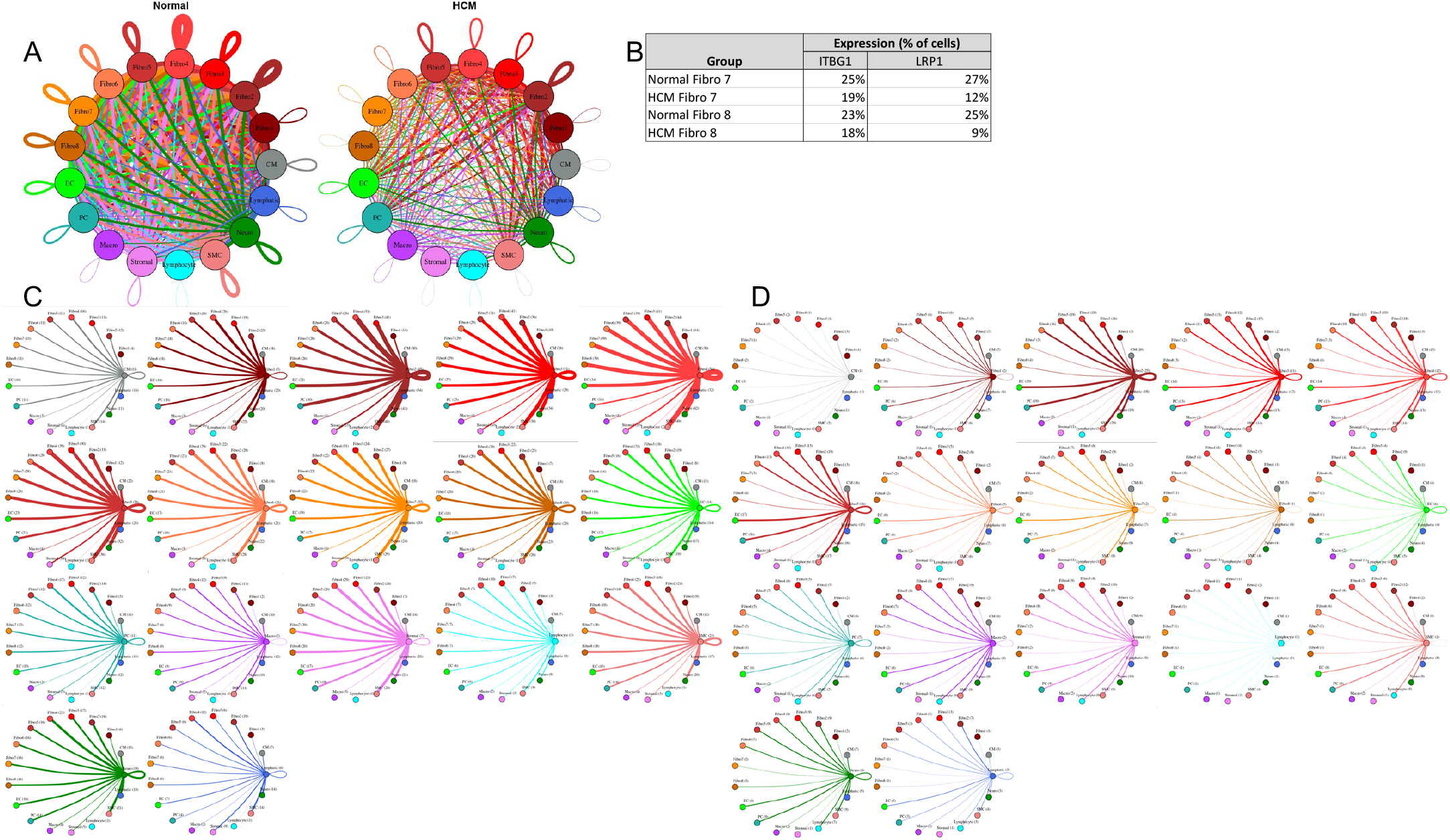
Cell-cell communication networks between fibroblast subtypes and other heart cells in normal control and HCM conditions. **A**. Comparison of Normal (left) and HCM (right) communication networks. Line color indicates ligand broadcast by the cell population with the same color. Lines connect to cell types which expressed cognate receptors. Line thickness is proportional to the number of uniquely expressed ligand-receptor pairs. Loops indicates communication within a cell type. **B**. Percent expression of receptors ITGB1 and LRP1 by fibroblast clusters (7 and 8) in normal and HCM conditions. **C, D**. Cell-cell communication networks broken down by cell type and fibroblast cluster in normal control (**C**) and HCM (**D**) conditions. Figure formatting follows panel **A**. Numbers indicate the quantity of uniquely expressed ligand-receptor pairs between the broadcasting cell type (expressing ligand) and receiving cell type (expressing receptor).

#### Cardiomyocyte subtypes and fibroblast subtypes together show cell subtype-specific alterations in cell-cell communication

Given the importance of sarcomere dysfunction in HCM and the observed reduction in ligand broadcasting of HCM cardiomyocytes (Fig. 2D), we examined the cell communication networks between fibroblast subtypes (i.e., clusters, n=8) and cardiomyocyte subtypes (n=13), in Normal and HCM tissue (Fig. 5). The total number of expressed ligand-receptor pairs in the Normal tissue was 2.6 times greater than in the HCM tissue (6975 and 2683 expressed ligand-receptor pairs respectively). Compared to the Normal tissue, HCM communication expectedly decreased drastically in most pathways (415 of 441 total pathways, Figure 5A-C). More so, 10.8% of pathways decreased to zero communication in HCM tissue. All zeroed communication pathways involved losses normally broadcasted from cardiomyocytes (clusters 2-5, 7, 11; Figure 5C, Supplemental Table ST5). Zeroed expression ligands (ligands that fell below the 20% expression threshold) in HCM cardiomyocytes included *TIMP1, COL1A2, COL3A1, FN1, LAMA2, COL1A2, CALM2, MFGE8, LPL, PSAP, SERPING1, COL6A1, COL6A2, VEGFA, SORBS1* and *ADAM17*. GO analysis of molecular functions associated with these ligands included platelet-derived growth factor binding, extracellular matrix structural constituent conferring tensile strength, collagen binding, protease binding and integrin binding (data not shown).

**Figure 5.**
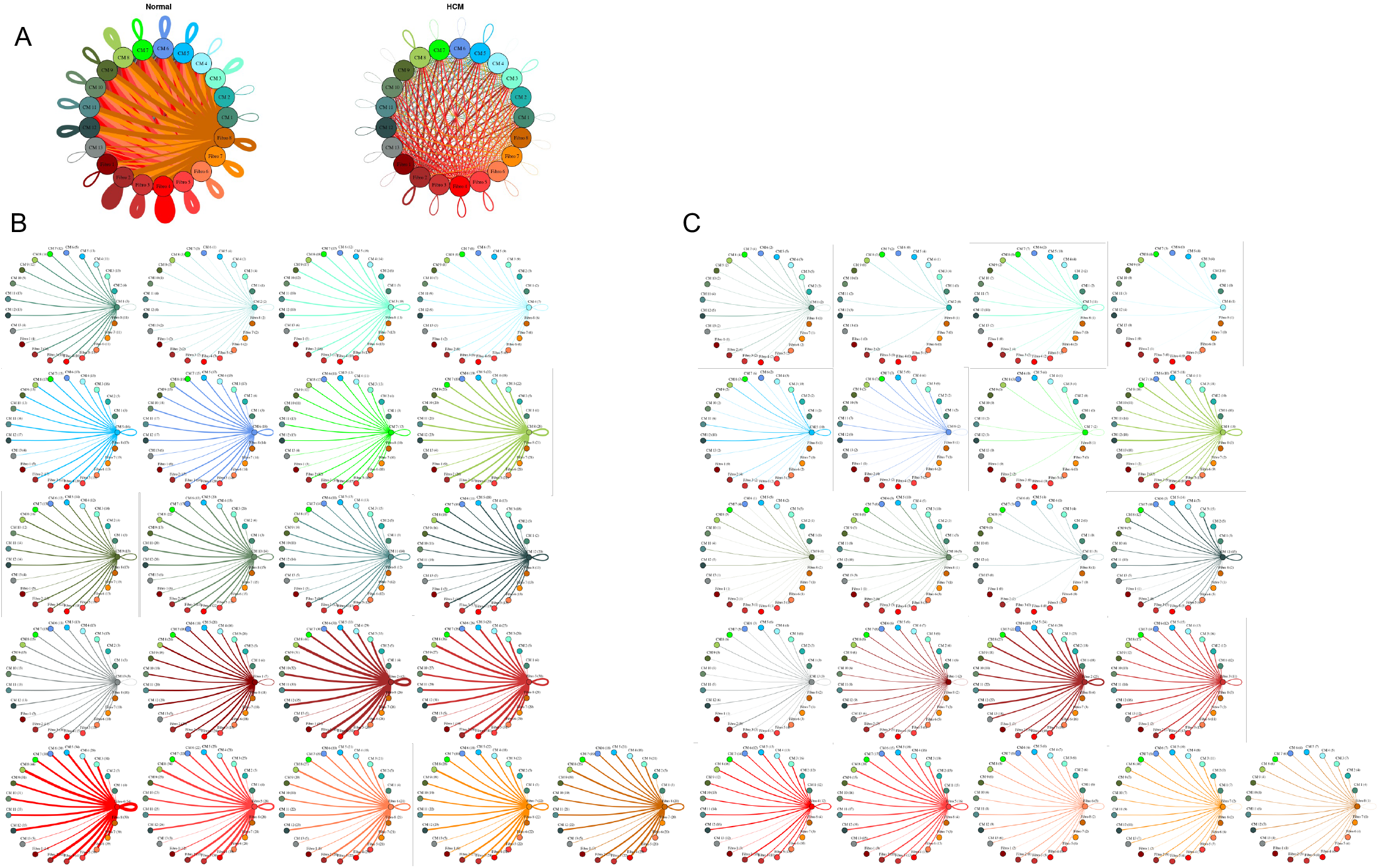
Cell-cell communication networks between cardiac fibroblast and cardiomyocyte subtypes in normal control and HCM conditions. **A**. Overall communication network between cardiomyocytes and fibroblasts. Line color indicates ligand broadcast by the cell population with the same color. Lines connect to cell types which expressed cognate receptors. Line thickness is proportional to the number of uniquely expressed ligand-receptor pairs. Loops indicates communication within a cell type. **B, C**. Cell-cell communication networks broken down by fibroblast cluster and cardiomyocyte cluster in normal control (**B**) and HCM (**C**) conditions. Numbers indicate the quantity of uniquely expressed ligand-receptor pairs between the broadcasting cell type (expressing ligand) and receiving cell type (expressing receptor).

The largest decreases in ligand-receptor pairs between Normal and HCM communication pathways (decreases >30 ligand-receptor pairs; Figure 4) involved fibroblast cluster 4 as fibroblast cluster 4 ligands were lost in HCM tissue compared to normal tissue (*C3 CALR, COL18A1, COL4A1, COL5A1, COL5A2, CXCL12, CYR61, FBLN1, FBN1, HSP90B1, IGF1, LAMA2, LAMB1, LAMC1, LGALS3BP, LRPAP1, NID1, PDGFD, PTN, TFPI, THBS2, TIMP2, TNC*; Supplemental Table ST6). The reduction in L-R pairs in fibroblast cluster 4 affected communication with other fibroblast clusters (Supplemental Figure ST7). The lost ligands are involved in vital molecular functions including integrin binding, extracellular matrix structural constituent, and extracellular matrix structural constituent conferring tensile strength among others, thereby suggesting fundamental alterations in these functions in HCM (Supplemental Table ST6).

While intercellular communication generally decreased in HCM tissue compared to Normal tissue, some specific cluster communication paths showed an opposite trend with increased HCM communication. HCM communication increased 3.2 times from fibroblasts clusters 2-5 to cardiomyocyte clusters 1, 2 and 13 compared to Normal tissue (Fig. 5c). This increase in potential communication can be attributed to 1) expression of the receptor ITGβ1 in cardiomyocyte clusters 1, 2 and 13 in HCM cells only, and 2) expression of several of ITGβ1’s cognate ligands in HCM fibroblast clusters (COL3A1, COL6A1, COL6A2, COL6A3, FBLN1, FBN1, FN1, HSPG2, LAMA2, LAMB1, LAMC1, LGALS3BP, TIMP2, VCAN). These cognate ligands were not expressed in Normal fibroblast clusters 2-5 (Supplemental Tables ST8). Another example of increased HCM communication (2.3 fold increase) relative to Normal communication involves communication from cardiomyocyte cluster 8 to cardiomyocyte clusters 1, 2 and 13 (Supplemental Table ST9).This increased communication can again be attributed to 1) expression of the of the receptor ITGβ1 in receiving clusters and 2) expression of several of ITGβ1’s cognate ligands in the broadcasting cardiomyocytes in cluster 8 (COL3A1, COL6A1, COL6A2, FN1, HSPG2, LAMA2, LGALS3BP). These cognate ligands were not expressed in Normal fibroblasts in cluster 8.

## Discussion

HCM is commonly referred to as a disease of the sarcomere, largely due to the number of sarcomere gene variants that have been identified in patients and families who carry the disease. Extensive progress has been made in determining how sarcomere mutations alter contractile function in cardiomyocytes. Such progress has led to therapies targeting sarcomere dysfunction ^27^. Little progress, however, has been made in understanding how these sarcomere mutations and other mutations causing sarcomere dysfunction result in the dysfunction of noncardiomyocyte cell types, although multiple nonsarcomeric mechanisms have been proposed ^28^. To address this gap in knowledge, we performed single nuclei RNA-sequencing and identified gene expression changes in various cell types from Normal IVS tissue and HCM myectomy tissue. We did not find groups of cells with distinctive gene expression patterns that could be labeled as an HCM-specific cell type, consequently supporting a hypothesis that subtle changes in gene expression within individual cell types likely promote the HCM phenotype.

Analysis of changes in cell-specific patterns of gene expression led to the identification of differentially expressed genes between Normal and HCM tissue. Differential expression analysis over UMAP space revealed several genes with different spatial and expression levels between Normal and HCM tissue, most notably sarcomere genes that exhibited increased expression in HCM and ECM genes that exhibited reduced expression in HCM. The changes in sarcomere gene expression may reflect compensatory changes due to sarcomere dysfunction or represent an increase associated with cardiac hypertrophy. The changes in ECM gene expression are consistent with qualitative changes in ECM properties in HCM hearts as has been previously reported ^29^. The observed alterations in ECM gene expression are reduced in HCM, however, which is paradoxical given the association of HCM with fibrosis. One possible explanation is that increased ECM matrix deposition may be associated with a steady state reduction in gene expression as protein accumulates, through a negative feedback loop.

Analysis of ligand-receptor pair gene expression has previously been used to infer a dense intercellular communication network in noncardiomyocyte cells of the normal mouse heart ^11^. This type of analysis, in which intercellular communication is thought to cease when expression is not detectable in more than 80% of the cells, is sensitive to subtle changes in ligand-receptor pair gene expression. Here we report that human HCM is associated with a general decrease in cell-specific expression of ligand-receptor pairs, regardless of genotype, a novel finding that suggests a common mechanism for how sarcomere dysfunction, which directly affects muscle cells, may also affect the function of non-muscle cell types. Specifically, the HCM phenotype is associated with a dramatic decrease in cardiac myocyte ligand broadcasting, with reduction in the ligands associated with interaction with the extracellular matrix. Concurrently, there is reduction in fibroblast ligand broadcasting, again with reduction in ligands associated with extracellular matrix interactions. These findings are highly suggestive that sarcomere dysfunction in HCM cardiomyocytes leads to abnormal interaction with the ECM, which in turn propagates further dysfunction through ECM signaling to cardiac fibroblasts, which also reduces further signaling through the ECM. Concurrently, relative to Normal tissue, there is increased expression of cognate ligands for the ITGβ1 receptor in subsets of cardiomyocytes and fibroblasts in HCM tissue that in turn activate a subset of cardiomyocytes and lymphocytes. These complex changes in cardiomyocyte to fibroblast signaling and fibroblast signaling to other cell types such as lymphocytes provide evidence for how sarcomere dysfunction in HCM propagates changes beyond individual cardiomyocytes. Changes in ECM, fibroblast, and lymphocyte function are particularly relevant to the development of myocyte disarray and fibrosis in HCM, with lymphocytes suggesting a potential inflammatory component. Within the IVS, fibroblasts are the most interactive cells compared to other cell types, consistent with a previous report in mice ^11^. In our dataset, fibroblasts interact extensively with smooth muscle cells, pericytes and endothelial cells which are all important components of the vasculature, and thus may explain the vascular abnormalities seen in HCM patients.

The most notable signaling pathways affected in HCM involve the receptor ITGβ1 and its cognate ligands, the majority of which are components of the ECM. ITGβ1 heterodimerizes with numerous α-integrins to form receptors for ECM molecules such as collagen, fibronectin and laminin. ITGβ1 is also localized to the costamere in cardiomyocytes to integrate both inside out and outside in signaling in mechanical signal transduction (reviewed in ^30^). Disruption of ITGβ1 function in the myocardium is associated with hypertrophy and fibrosis ^31^, but a role in HCM has not yet been described, to the best of our knowledge. The cardiac ECM is dynamic throughout development and after myocardial injury, with components affecting the function of cardiac cells and determining the stiffness of cardiac tissue. In a pig model of HCM carrying a MYH7^R403Q^ variant, the ECM has been demonstrated to promote impaired relaxation, increased force development, and increased stiffness in wild type human iPSC-derived cardiomyocytes. These results indicate that the HCM ECM is functionally altered as a result of sarcomere dysfunction, and likely reinforces the contractile alterations in cardiomyocytes that result from sarcomere mutations ^29^. Our results showing altered intercellular communication between cardiomyocytes, fibroblasts, and other cell types, through changes in ECM component expression and signaling, provide a conceptual basis for how an HCM-specific ECM may be generated.

The importance of sarcomere mutations has been widely recognized but lack of information regarding phenotypic features that cannot be easily explained by these mutations is also a commonly recognized gap in existing knowledge, leading some to postulate interactions between cardiomyocytes and noncardiomyocytes as a potential cause ^32, 33^. Others have found that treatment focusing on sarcomere gene dysfunction can improve sarcomere function but do not promote regression of hypertrophy and fibrosis in animal models ^27, 34^. Animal models of HCM have important differences from human HCM ^35^, however, and thus future discovery work for developing therapies must involve either human tissue or high-fidelity models. A recent study of HCM patients with extreme fibrotic phenotypes has implicated JAK2 as a potential contributor, for example ^36^. Our study is the first to document altered L-R gene expression at the single cell level that suggests altered intercellular communication and ECM signaling in samples derived from human patients, mostly lacking sarcomere gene mutations but with clinically documented, symptomatic obstructive HCM. Population-based studies have suggested that HCM patients without sarcomere gene mutations have a better prognosis ^37^, leading others to suggest that sarcomere mutation negative HCM is a complex polygenic disorder ^7^. Further delineation of the intercellular cross talk in human HCM and the specific signals that promote adverse ECM remodeling combined with therapies targeting the sarcomere will likely lead to further improvement in the treatment of human HCM.

The clinical implications of this work are that another paradigm for HCM pathogenesis beyond sarcomere dysfunction needs to be considered, involving intercellular communication between cardiomyocytes, the extracellular matrix, fibroblasts and components of the immune system. Understanding the roles of cells other than myocytes will identify other potential therapeutic targets for disease-specific and personalized therapies. The discovery of a potential role for integrin-β1 and its ECM ligands in human HCM-associated LVOT obstruction provides an opportunity for further mechanistic studies in experimental models and also for potential clinical trials with FDA-approved anti-integrin therapies. A limitation of our study is that single nuclei RNA-sequencing does not necessarily reflect changes in protein expression, and thus L-R protein expression at the single cell level may not directly correlate with our findings.

## Data Availability

All data are available in the Gene Expression Omnibus database under accession numbers GSE161921 and GSE174691

https://www.ncbi.nlm.nih.gov/geo

## Abbreviations

HCM: Hypertrophic Cardiomyopathy
snRNA-seq: Single nuclei RNA-sequencing
ECM: Extracellular Matrix
LVOT: Left Ventricular Outflow Tract
IVS: Interventricular Septum
L-R: Ligand-Receptor
ITGβ1: Integrin-β1
QC: Quality Control
UMAP: Uniform Manifold Approximation and Projection
GO: Gene Ontology
TGF-β: Transforming Growth Factor-β

