## supplemental tables and figures for "Altered Intercellular Communication and Extracellular Matrix Signaling as a Potential Disease Mechanism in Human Hypertrophic Cardiomyopathy"

### Supplemental Table ST1. Patient Characteristics.

| Patient | 1 | 2 | 3 | 4 | 5 | 6 | 7 | 8 | 9 |
|---|---|---|---|---|---|---|---|---|---|
| Demographics | | | | | | | | | |
| Age range at myectomy | 41-45 | 51-55 | 51-55 | 51-55 | 71-75 | 56-60 | 51-55 | 61-65 | 56-60 |
| female | no | yes | yes | no | yes | no | no | no | no |
| nyha class>3 | yes | yes | yes | yes | yes | yes | no | yes | yes |
| Med Hx | | | | | | | | | |
| Prior AF | no | no | no | yes | no | no | no | yes | no |
| Prior VT/VF | no | no | no | no | no | no | no | no | no |
| Prior NS VT | no | no | no | no | no | no | no | no | no |
| Prior syncope | no | no | no | no | no | no | yes | no | no |
| Fam Hx SCD | no | no | yes | no | no | no | no | no | no |
| Fam Hx HCM | no | yes | yes | no | no | no | no | no | no |
| Comorbidities | none | CAD, HTN, HLD, COPD, DM2, OSA, Spinal Stenosis | HTN, HLD | HTN | OSA, HTN, CAD, HLD | HTN, HLD, CAD | none | HLD | prostate CA, OSA |
| Meds | | | | | | | | | |
| beta blocker | yes | yes | yes | yes | no | yes | no | yes | yes |
| calcium channel blocker | no | no | no | yes | yes | no | yes | no | no |
| ACE or ARB | no | no | no | yes | no | no | no | no | no |
| Diuretic Use | no | no | no | no | no | yes | no | no | no |
| loop diuretic | no | no | no | no | no | yes | no | no | no |
| thiazide | no | no | no | no | no | no | no | no | no |
| potassium sparing | no | no | no | no | no | no | no | no | no |
| disopyramide | no | no | no | no | no | no | no | no | no |
| amiodarone | no | no | no | no | no | no | no | no | no |
| Physiological measurements | | | | | | | | | |
| LA size (mm) | 52 | 46 | 40 | 69 | 49 | 54 | 35 | 57 | 39 |
| systolic blood pressure | 128 | 110 | 106 | 124 | 126 | 142 | 140 | 126 | 140 |
| diastolic blood pressure | 82 | 80 | 60 | 78 | 78 | 90 | 80 | 78 | 80 |
| IVS thickness (mm) | 13 | 15 | 22 | 24 | 15 | 15 | 15 | 18 | 18 |
| Posterior wall thickness | 12 | 12 | 13 | 11 | 8.9 | 8.7 | 9.6 | 14 | 13 |
| LVEF (%) | 70 | 65 | 65 | 60 | 65 | 65-70 | 65 | 65 | 70 |
| LVEDD (mm) | 45 | 36 | 36 | 43 | 31 | 48 | 40 | 44 | 33 |
| LVESD (m) | 29 | 23 | 22 | 33 | 22 | 33 | 24 | 25 | 21 |
| SAM | yes | yes | yes | yes | yes | yes | yes | yes | no |
| MR | moderate | mild | mild | moderate to severe | mild | trace | mild | trace | mild |
| LVOT gradient rest (mm Hg) | 60 | 35 | 110 | 0 | 0 | 0 | 75 | 100 | 0 |
| LVOT gradient provocation (mm Hg) | NA | 85 | NA | 40 | 150 | 110 | NA | NA | 90-100 |
| LGE on MRI | none | mild | ND | moderate | none | none | ND | none | mild |
| Surgical Procedure | extended septal myectomy with mitral valve repair | extended septal myectomy, CABGx1 | extended septal myectomy | extended septal myectomy, MV repair, MAZE | extended septal myectomy | septal myectomy and MV repair | extended septal myectomy | extended septal myectomy, MAZE | extended septal myectomy |
| Pathogenic HCM Variant | NF | NF | NF | KRAS | NF | NF | NF | NF | MYBPC3 |

Abbreviations: NYHA = New York Heart Association; AF = atrial fibrillation; VT/VF = ventricular tachycardia or ventricular fibrillation; NSVT = nonsustained ventricular tachycardia; SCD = sudden cardiac death; HCM = hypertrophic cardiomyopathy; CAD = coronary artery disease; HTN = hypertension; HLD = hyperlipidemia; COPD = chronic obstructive pulmonary disease; DM2 = diabetes mellitus, type 2; OSA = obstructive sleep apnea; DI = diabetes insipidus; GERD = gastroesophageal reflux disease; AS = aortic stenosis; MR = mitral regurgitation; NF = not found; ND = not done; ICD = implantable cardioverter-defibrillator

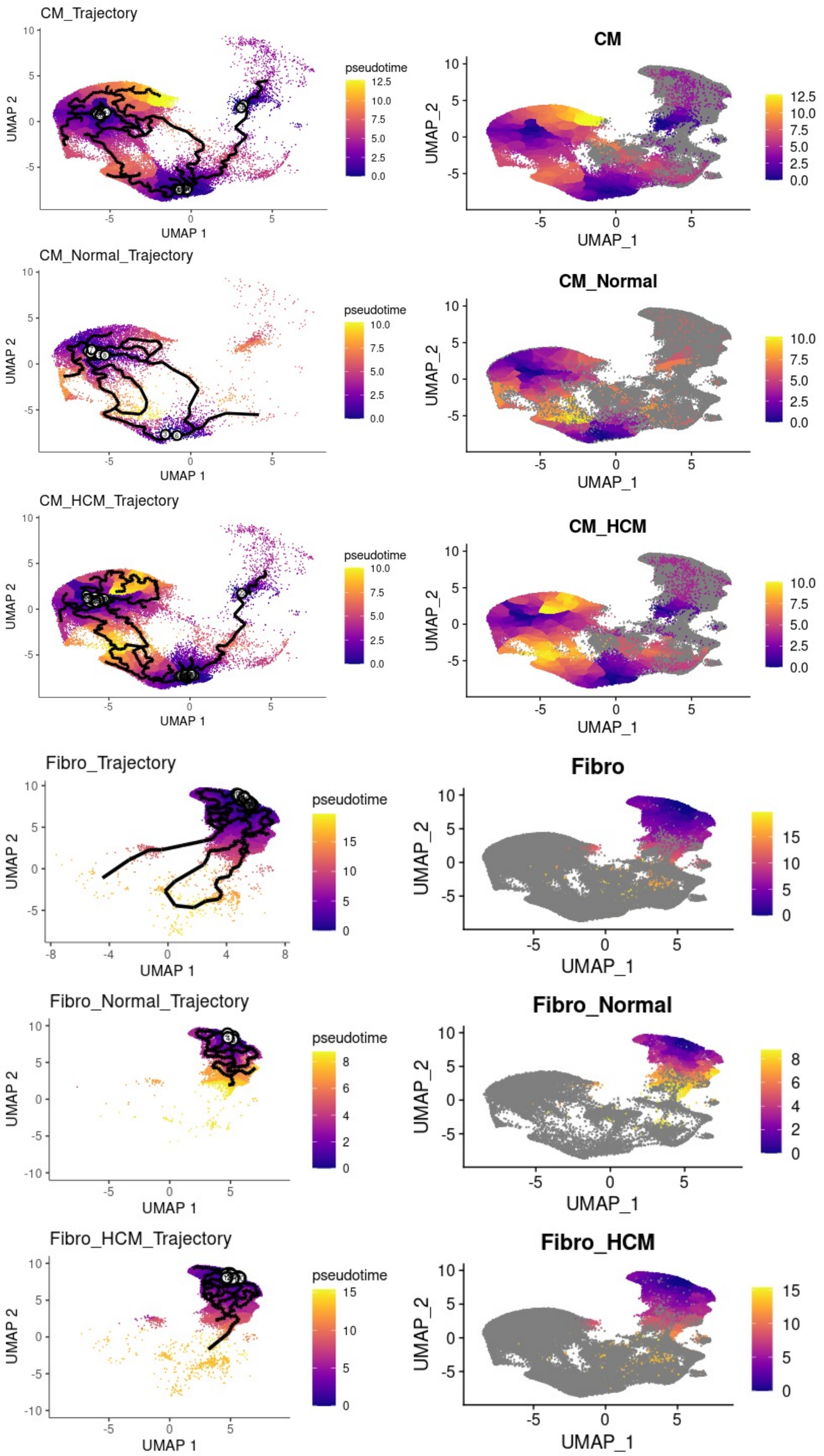

**Supplemental Figure S1**. Trajectory analysis of cardiomyocytes and fibroblasts in normal and HCM heart tissue. Trajectories are similar in normal and HCM cardiomyocytes and in normal and HCM fibroblasts.

### Supplemental Table ST2. Number of Differentially Expressed Genes for Each Cell Type, Along Trajectory, Determined by Spatial Autocorrelation

| Cell Type | Cell Class | Number of Cells | Number of Differentially Expressed Genes | Number of Gene Overlap |
|---|---|---|---|---|
| CM | Normal | 10131 | 2818 | 1823 |
| | HCM | 57847 | 3589 | |
| Fibro | Normal | 6935 | 573 | 300 |
| | HCM | 22830 | 481 | |
| EC | Normal | 1526 | 661 | 295 |
| | HCM | 9571 | 614 | |
| PC | Normal | 1010 | 320 | 127 |
| | HCM | 6310 | 482 | |
| Macro | Normal | 2734 | 278 | 129 |
| | HCM | 5821 | 402 | |
| Lymphocyte | Normal | 300 | 325 | 130 |
| | HCM | 922 | 358 | |
| Stromal | Normal | 1350 | 94 | 28 |
| | HCM | 3895 | 91 | |
| SMC | Normal | 511 | 547 | 173 |
| | HCM | 1498 | 492 | |
| Neuro | Normal | 188 | 114 | 6 |
| | HCM | 632 | 137 | |
| Lymphatic | Normal | 173 | 105 | 4 |
| | HCM | 497 | 148 | |

### Supplemental Table ST3. Differentially Expressed Genes Along Trajectories, Determined by Spatial Autocorrelation, Listed Alphabetically with Cell Type

| | Filtered Differentially Expressed Genes Over Space | Cell Type |
|---|---|---|
| 1 | ABCA10 | Endothelial |
| 2 | ABCA6 | Cardiomyocyte, Pericyte |
| 3 | ABCA8 | Cardiomyocyte |
| 4 | ABLIM1 | Lymphatic |
| 5 | ACTA1 | Neuronal |
| 6 | ACTC1 | Cardiomyocyte, Lymphatic |
| 7 | ACTG1 | Smooth Muscle |
| 8 | ADH1B | Cardiomyocyte, Nueronal |
| 9 | ADJ1B | Pericyte |
| 10 | AGT | Endothelial, AGT |
| 11 | AKAP13 | Lymphatic |
| 12 | ALDOA | Smooth Muscle |
| 13 | ANKRD1 | Lymphatic |
| 14 | ANKRD2 | Smooth Muscle, Lymphatic |
| 15 | APOE | Fibroblast |
| 16 | AQP7 | Macrophage |
| 17 | ASPM | Endothelial |
| 18 | ATP1A2 | Lymphatic |
| 19 | B2M | Macrophage, Lymphocyte |
| 20 | C1R | Fibroblast |
| 21 | C1S | Fibroblast |
| 22 | C7 | Fibroblast |
| 23 | CCL2 | Neuronal |
| 24 | CCL21 | Lymphatic |
| 25 | CD2 | Lymphocyte |
| 26 | CD36 | Smooth Muscle, Lymphatic |
| 27 | CD63 | Smooth Muscle |
| 28 | CDH19 | Neuronal |
| 29 | CFD | Endothelial |
| 30 | CFL1 | Endothelial |
| 31 | CILP | Fibroblast |
| 32 | CMC2 | Neuronal |
| 33 | CMYA5 | Cardiomyocyte |
| 34 | COA7 | Lymphatic |
| 35 | COL1A1 | Fibroblast |
| 36 | COL6A1 | Fibroblast |
| 37 | COL6A2 | Fibroblast |
| 38 | COX6A2 | Lymphatic |
| 39 | CPE | Endothelial |
| 40 | CSDC2 | Neuronal |
| 41 | CTBS | Lymphatic |
| 42 | CYR61 | Fibroblast |
| 43 | DCN | Cardiomyocyte, Endothelial |
| 44 | DCP2 | Cardiomyocyte |
| 45 | DES | Cardiomyocyte |
| 46 | DNAJC8 | Neuronal |
| 47 | DNM3OS | Pericyte |
| 48 | DNUFA5 | Lymphatic |
| 49 | EEF1A1 | Endothelial, Lymphatic |
| 50 | EHMT1 | Lymphatic |
| 51 | EXOG | Smooth Muscle |
| 52 | FABP4 | Smooth Muscle, Lymphatic |
| 53 | FAM134B | Lymphatic |
| 54 | FHL2 | Cardiomyocyte, Lympahtic |
| 55 | FLOT1 | Neuronal |
| 56 | FOS | Cardiomyocyte, Macrophage |
| 57 | FTH1 | Endothelial |
| 58 | GAPDH | Smooth Muscle |
| 59 | GAS7 | Lymphatic |
| 60 | GJA5 | Endothelial |
| 61 | GPNMB | Neuronal |
| 62 | GPR155 | Cardiomyocyte |
| 63 | GPRIN3 | Cardiomyocyte |
| 64 | GRHPR | Neuronal |
| 65 | GZMK | Lymphocyte |
| 66 | H1FX | Neuronal |
| 67 | H3F3B | Smooth Muscle |
| 68 | HES1 | Fibroblast |
| 69 | HIC1 | Smooth Muscle |
| 70 | HLA-B | Smooth Muscle |
| 71 | HLA-DPB1 | Macrophage |
| 72 | HLA-DRA | Macrophage, Lymphocyte |
| 73 | HMGN1 | Lymphatic |
| 74 | HSF4 | Smooth Muscle |
| 75 | ID4 | Smooth Muscle |
| 76 | IDGFBP7 | Lymphatic |
| 77 | IGF2 | Smooth Muscle |
| 78 | IGFBP3 | Endothelial |
| 79 | IGFBP7 | Fibroblast |
| 80 | INHBA | Endothelial |
| 81 | IRS1 | Neuronal |
| 82 | JUNB | Cardiomyocyte |
| 83 | KCNJ8 | Smooth Muscle |
| 84 | KIF1C | Cardiomyocyte |
| 85 | LEPR | Endothelial |
| 86 | LGALS1 | Smooth Muscle |
| 87 | LINC01089 | Neuronal |
| 88 | MALAT1 | Cardiomyocyte, Fibroblast, Endothelial, Macrophage, Lymphocyte, Smooth Muscle, Neuronal |
| 89 | MEG3 | Cardiomyocyte, Macrophage |
| 90 | MFAP4 | Fibroblast |
| 91 | MFSD14A | Neuronal |
| 92 | MPC1 | Lymphatic |
| 93 | MPHOSPH8 | Neuronal |
| 94 | MRRF | Lymphatic |
| 95 | MS4A6A | Cardiomyocyte |
| 96 | MYH7 | Cardiomyocyte |
| 97 | MYH7B | Macrophage |
| 98 | MYL2 | Cardiomyocyte, Smooth Muscle, Neuronal |
| 99 | MZT2A | Lymphatic |
| 100 | NDUFA1 | Neuronal |
| 101 | NDUFB2 | Neuronal |
| 102 | NEAT1 | Cardiomyocyte, Pericyte |
| 103 | NEBL | Macrophage |
| 104 | NMRK2 | Lymphatic |
| 105 | NOVA1 | Neuronal |
| 106 | NPPA | Cardiomyocyte |
| 107 | NPPB | Cardiomyocyte |
| 108 | PDGFRB | Macrophage |
| 109 | PDLIM5 | Lymphatic |
| 110 | PGM3 | Neuronal |
| 111 | PKN2 | Neuronal |
| 112 | PLA2G2A | Cardiomyocyte, Endothelial |
| 113 | PMEPA1 | Neuronal |
| 114 | POLR2J3 | Neuronal |
| 115 | POSTN | Fibroblast, Endothelial |
| 116 | PPHLN1 | Neuronal |
| 117 | PPRG4 | Fibroblast |
| 118 | PRKDC | Neuronal |
| 119 | PTMA | Smooth Muscle |
| 120 | PTPRS | Smooth Muscle |
| 121 | RAB8B | Smooth Muscle |
| 122 | RBFOX2 | Neuronal |
| 123 | RCN2 | Cardiomyocyte |
| 124 | RMND5B | Smooth Muscle |
| 125 | RP11-39404.5 | Smooth Muscle |
| 126 | RP11-983P16.4 | Neuronal |
| 127 | RPL10 | Smooth Muscle |
| 128 | RPL11 | Lymphatic |
| 129 | RPL13A | Endothelial |
| 130 | RPL23A | Endothelial |
| 131 | RPL3 | Endothelial |
| 132 | RPL7 | Endothelial |
| 133 | RPS15 | Lymphatic |
| 134 | RRP15 | Lymphatic |
| 135 | S100A1 | Smooth Muscle |
| 136 | SAT1 | Cardiomyocyte |
| 137 | SDHA | Lymphatic |
| 138 | SEPHS1 | Neuronal |
| 139 | SERINC3 | Lymphatic |
| 140 | SERPINF1 | Cardiomyocyte, Fibroblast |
| 141 | SF1 | Neuronal |
| 142 | SLC11A2 | Neuronal |
| 143 | SLC25A6 | Smooth Muscle |
| 144 | SLC35F6 | Neuronal |
| 145 | SLC41A1 | Lymphatic |
| 146 | SLCO5A1 | Neuronal |
| 147 | SLMAP | Pericyte |
| 148 | SMIM8 | Smooth Muscle |
| 149 | SMU1 | Lymphatic |
| 150 | SNHG21 | Neuronal |
| 151 | SORBS2 | Smooth Muscle, Lymphatic |
| 152 | SREK1 | Lymphatic |
| 153 | STAT6 | Cardiomyocyte |
| 154 | STEAP4 | Smooth Muscle |
| 155 | SYNPO | Cardiomyocyte |
| 156 | TAGLN | Smooth Muscle |
| 157 | TECRL | Smooth Muscle |
| 158 | TFPI | Smooth Muscle |
| 159 | TGFB2 | Lymphatic |
| 160 | TMED4 | Cardiomyocyte |
| 161 | TMEM175 | Neuronal |
| 162 | TMSB4X | Endothelial, Smooth Muscle |
| 163 | TNNI3 | Cardiomyocyte, Smooth Muscle |
| 164 | TNNT2 | Cardiomyocyte, Lympahtic |
| 165 | TPCN1 | Neuronal |
| 166 | TRMT112 | Lymphatic |
| 167 | TUBA1B | Smooth Muscle |
| 168 | TYROBP | Neuronal |
| 169 | UBR3 | Neuronal |
| 170 | UGDH-AS1 | Neuronal |
| 171 | UTRN | Lymphatic |
| 172 | WASF2 | Neuronal |
| 173 | YAP1 | Neuronal |
| 174 | ZMYM2 | Cardiomyocyte |
| 175 | ZNF106 | Cardiomyocyte |
| 176 | ZNF211 | Lymphatic |

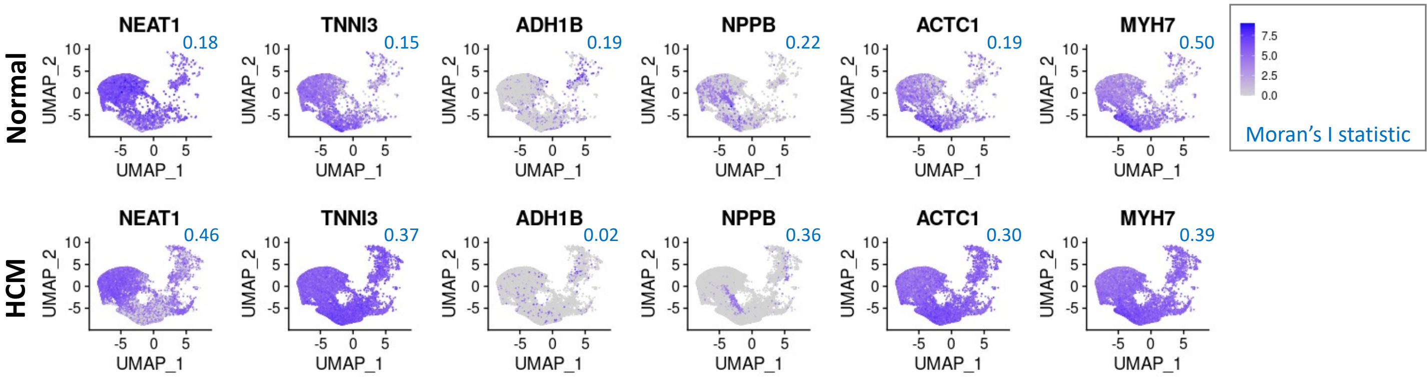

**Supplemental Figure S2**. Selected cardiomyocyte genes that are differentially expressed along the cardiomyocyte trajectory by spatial autocorrelation, represented in UMAP space for Normal and HCM cells

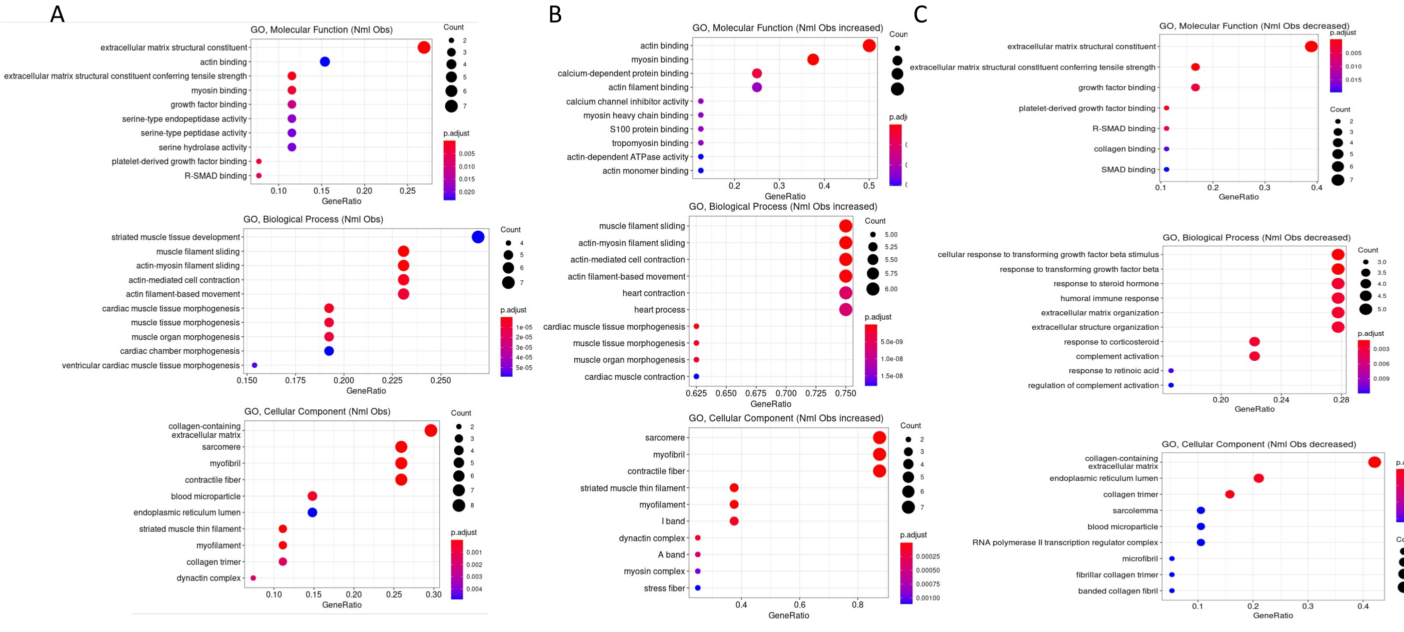

Supplemental Figure S3. Gene Ontology Enrichment Analysis of Differentially Expressed Genes in HCM. A. GO enrichment analysis of all differentially expressed genes in HCM from Table 1. B. GO enrichment analysis of Table 1 genes increased in HCM. C. GO enrichment analysis of Table 1 genes decreased in HCM.

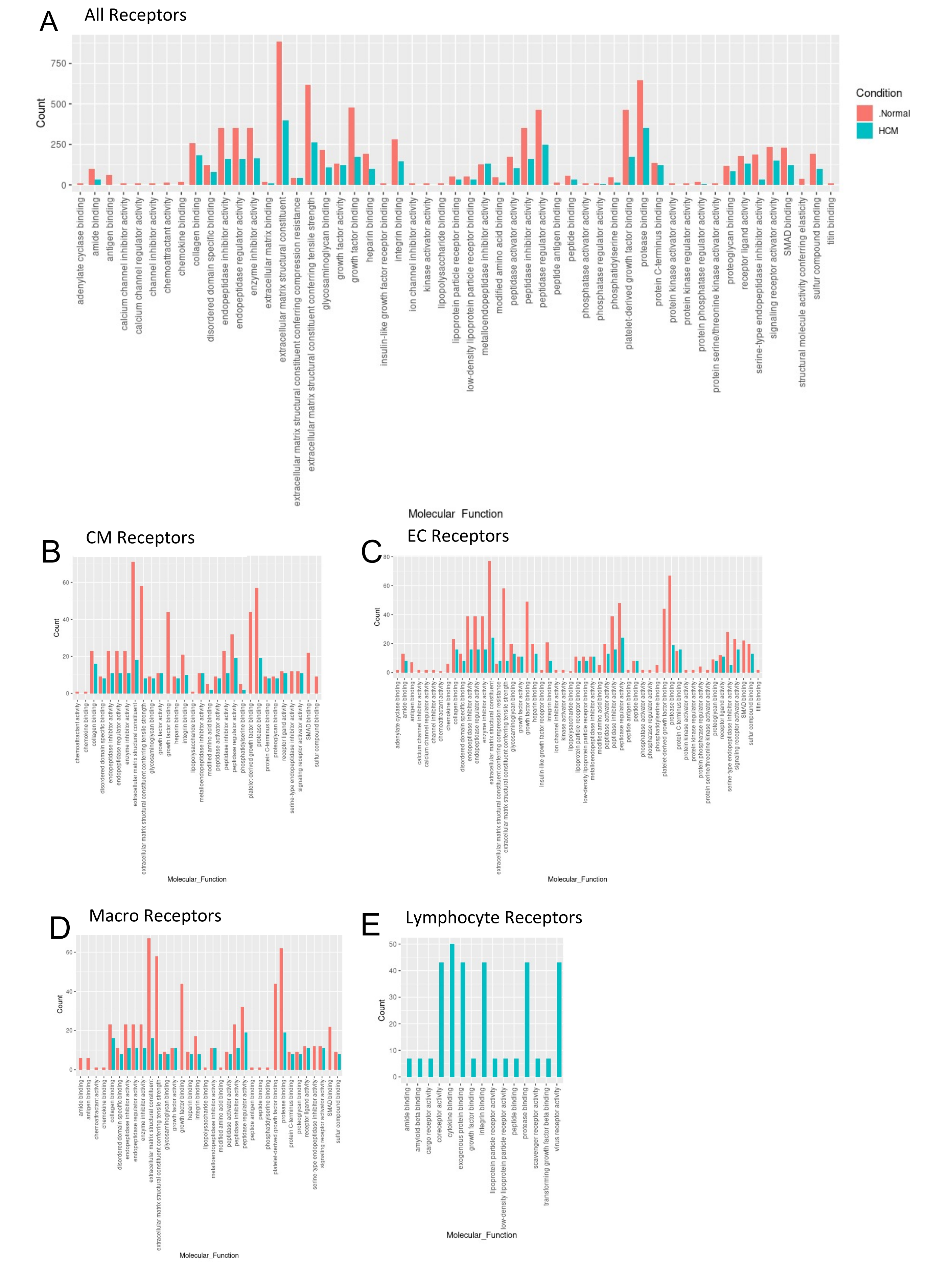

**Supplemental Figure S4.** Bar plot representing the total count of receptors (in expressed ligand-receptor pairs) associated with different cellular processes in Normal and HCM IVS Cells. Bar color distinguishes ligand count in normal or HCM conditions. A. Comparison of molecular processes across all cell types. B. Comparison in cardiomyocytes. C. Endothelial cells. D. Macrophages. E. Lymphocytes

### Supplemental Table ST4: Fibroblast to Lymphocyte Ligand-Receptor Interactions in Normal and HCM IVS Tissue

| **Normal Condition** | | | | | | **HCM Condition** | | | | |
|---|---|---|---|---|---|---|---|---|---|---|
| **L-Cell** | **R-Cell** | **L** | **R** | **L-R pair** | | **L-Cell** | **R-Cell** | **L** | **R** | **L-R pair** |
| Fibro | Lymphocyte | TIMP1 | CD63 | TIMP1_CD63 | | Fibro | Lymphocyte | COL1A1 | CD36 | COL1A1_CD36 |
| | | | | | | Fibro | Lymphocyte | COL1A2 | CD36 | COL1A2_CD36 |
| | | | | | | Fibro | Lymphocyte | COL1A1 | ITGB1 | COL1A1_ITGB1 |
| | | | | | | Fibro | Lymphocyte | COL1A2 | ITGB1 | COL1A2_ITGB1 |
| | | | | | | Fibro | Lymphocyte | COL3A1 | ITGB1 | COL3A1_ITGB1 |
| | | | | | | Fibro | Lymphocyte | COL6A1 | ITGB1 | COL6A1_ITGB1 |
| | | | | | | Fibro | Lymphocyte | COL6A2 | ITGB1 | COL6A2_ITGB1 |
| | | | | | | Fibro | Lymphocyte | COL6A3 | ITGB1 | COL6A3_ITGB1 |
| | | | | | | Fibro | Lymphocyte | FBLN1 | ITGB1 | FBLN1_ITGB1 |
| | | | | | | Fibro | Lymphocyte | FN1 | ITGB1 | FN1_ITGB1 |
| | | | | | | Fibro | Lymphocyte | HSPG2 | ITGB1 | HSPG2_ITGB1 |
| | | | | | | Fibro | Lymphocyte | VCAN | ITGB1 | VCAN_ITGB1 |
| | | | | | | Fibro | Lymphocyte | TIMP1 | CD63 | TIMP1_CD63 |

### Supplemental Table ST5, Cardiomyocyte Communication Pathways No Longer Present in HCM

| L-Cell | R-Cell | L | R | L-R pair |
|---|---|---|---|---|
| CM 2 | CM 1 | TIMP1 | CD63 | TIMP1_CD63 |
| CM 2 | CM 2 | TIMP1 | CD63 | TIMP1_CD63 |
| CM 2 | CM 2 | CALM2 | INSR | CALM2_INSR |
| CM 2 | CM 6 | TIMP1 | CD63 | TIMP1_CD63 |
| CM 2 | CM 9 | TIMP1 | CD63 | TIMP1_CD63 |
| CM 2 | CM 9 | CALM2 | INSR | CALM2_INSR |
| CM 2 | CM 10 | TIMP1 | CD63 | TIMP1_CD63 |
| CM 2 | Fibro 1 | TIMP1 | CD63 | TIMP1_CD63 |
| CM 2 | Fibro 1 | MFGE8 | PDGFRB | MFGE8_PDGFRB |
| CM 2 | Fibro 3 | TIMP1 | CD63 | TIMP1_CD63 |
| CM 2 | Fibro 3 | MFGE8 | PDGFRB | MFGE8_PDGFRB |
| CM 2 | Fibro 4 | TIMP1 | CD63 | TIMP1_CD63 |
| CM 2 | Fibro 4 | MFGE8 | PDGFRB | MFGE8_PDGFRB |
| CM 2 | Fibro 4 | MFGE8 | ITGAV | MFGE8_ITGAV |
| CM 2 | Fibro 6 | TIMP1 | CD63 | TIMP1_CD63 |
| CM 2 | Fibro 6 | MFGE8 | PDGFRB | MFGE8_PDGFRB |
| CM 2 | Fibro 7 | TIMP1 | CD63 | TIMP1_CD63 |
| CM 2 | Fibro 7 | MFGE8 | PDGFRB | MFGE8_PDGFRB |
| CM 2 | CM 13 | TIMP1 | CD63 | TIMP1_CD63 |
| CM 2 | CM 13 | CALM2 | INSR | CALM2_INSR |
| CM 3 | Fibro 1 | MFGE8 | PDGFRB | MFGE8_PDGFRB |
| CM 3 | Fibro 1 | TIMP1 | CD63 | TIMP1_CD63 |
| CM 3 | Fibro 1 | LPL | LRP1 | LPL_LRP1 |
| CM 3 | Fibro 1 | PSAP | LRP1 | PSAP_LRP1 |
| CM 3 | Fibro 1 | SERPING1 | LRP1 | SERPING1_LRP1 |
| CM 3 | Fibro 7 | MFGE8 | PDGFRB | MFGE8_PDGFRB |
| CM 3 | Fibro 7 | TIMP1 | CD63 | TIMP1_CD63 |
| CM 3 | Fibro 7 | LPL | LRP1 | LPL_LRP1 |
| CM 3 | Fibro 7 | PSAP | LRP1 | PSAP_LRP1 |
| CM 3 | Fibro 7 | SERPING1 | LRP1 | SERPING1_LRP1 |
| CM 3 | Fibro 7 | COL1A1 | ITGB1 | COL1A1_ITGB1 |
| CM 3 | Fibro 7 | COL1A2 | ITGB1 | COL1A2_ITGB1 |
| CM 3 | Fibro 7 | COL3A1 | ITGB1 | COL3A1_ITGB1 |
| CM 3 | Fibro 7 | COL6A1 | ITGB1 | COL6A1_ITGB1 |
| CM 3 | Fibro 7 | COL6A2 | ITGB1 | COL6A2_ITGB1 |
| CM 3 | Fibro 7 | FN1 | ITGB1 | FN1_ITGB1 |
| CM 3 | Fibro 7 | LAMA2 | ITGB1 | LAMA2_ITGB1 |
| CM 4 | CM 1 | TIMP1 | CD63 | TIMP1_CD63 |
| CM 4 | CM 1 | COL1A2 | CD36 | COL1A2_CD36 |
| CM 4 | CM 2 | TIMP1 | CD63 | TIMP1_CD63 |
| CM 4 | CM 2 | COL1A2 | CD36 | COL1A2_CD36 |
| CM 4 | CM 2 | CALM2 | INSR | CALM2_INSR |
| CM 4 | CM 6 | COL1A2 | ITGB1 | COL1A2_ITGB1 |
| CM 4 | CM 6 | COL3A1 | ITGB1 | COL3A1_ITGB1 |
| CM 4 | CM 6 | FN1 | ITGB1 | FN1_ITGB1 |
| CM 4 | CM 6 | LAMA2 | ITGB1 | LAMA2_ITGB1 |
| CM 4 | CM 6 | TIMP1 | CD63 | TIMP1_CD63 |
| CM 4 | CM 6 | COL1A2 | CD36 | COL1A2_CD36 |
| CM 4 | CM 6 | LAMA2 | RPSA | LAMA2_RPSA |
| CM 4 | CM 9 | TIMP1 | CD63 | TIMP1_CD63 |
| CM 4 | CM 9 | COL1A2 | CD36 | COL1A2_CD36 |
| CM 4 | CM 9 | CALM2 | INSR | CALM2_INSR |
| CM 4 | CM 9 | COL1A2 | ITGB1 | COL1A2_ITGB1 |
| CM 4 | CM 9 | COL3A1 | ITGB1 | COL3A1_ITGB1 |
| CM 4 | CM 9 | FN1 | ITGB1 | FN1_ITGB1 |
| CM 4 | CM 9 | LAMA2 | ITGB1 | LAMA2_ITGB1 |
| CM 4 | CM 9 | CALM2 | CACNA1C | CALM2_CACNA1C |
| CM 4 | CM 10 | COL1A2 | ITGB1 | COL1A2_ITGB1 |
| CM 4 | CM 10 | COL3A1 | ITGB1 | COL3A1_ITGB1 |
| CM 4 | CM 10 | FN1 | ITGB1 | FN1_ITGB1 |
| CM 4 | CM 10 | LAMA2 | ITGB1 | LAMA2_ITGB1 |
| CM 4 | CM 10 | TIMP1 | CD63 | TIMP1_CD63 |
| CM 4 | CM 10 | COL1A2 | CD36 | COL1A2_CD36 |
| CM 4 | CM 10 | LAMA2 | RPSA | LAMA2_RPSA |
| CM 4 | Fibro 1 | MFGE8 | PDGFRB | MFGE8_PDGFRB |
| CM 4 | Fibro 1 | TIMP1 | CD63 | TIMP1_CD63 |
| CM 4 | Fibro 3 | MFGE8 | PDGFRB | MFGE8_PDGFRB |
| CM 4 | Fibro 3 | TIMP1 | CD63 | TIMP1_CD63 |
| CM 4 | Fibro 3 | COL1A2 | ITGB1 | COL1A2_ITGB1 |
| CM 4 | Fibro 3 | COL3A1 | ITGB1 | COL3A1_ITGB1 |
| CM 4 | Fibro 3 | FN1 | ITGB1 | FN1_ITGB1 |
| CM 4 | Fibro 3 | LAMA2 | ITGB1 | LAMA2_ITGB1 |
| CM 4 | Fibro 4 | TIMP1 | CD63 | TIMP1_CD63 |
| CM 4 | Fibro 4 | COL1A2 | ITGB1 | COL1A2_ITGB1 |
| CM 4 | Fibro 4 | COL3A1 | ITGB1 | COL3A1_ITGB1 |
| CM 4 | Fibro 4 | FN1 | ITGB1 | FN1_ITGB1 |
| CM 4 | Fibro 4 | LAMA2 | ITGB1 | LAMA2_ITGB1 |
| CM 4 | Fibro 4 | FN1 | SDC2 | FN1_SDC2 |
| CM 4 | Fibro 4 | COL1A2 | ITGAV | COL1A2_ITGAV |
| CM 4 | Fibro 4 | FN1 | ITGAV | FN1_ITGAV |
| CM 4 | Fibro 4 | MFGE8 | ITGAV | MFGE8_ITGAV |
| CM 4 | Fibro 6 | MFGE8 | PDGFRB | MFGE8_PDGFRB |
| CM 4 | Fibro 6 | TIMP1 | CD63 | TIMP1_CD63 |
| CM 4 | Fibro 6 | COL1A2 | ITGB1 | COL1A2_ITGB1 |
| CM 4 | Fibro 6 | COL3A1 | ITGB1 | COL3A1_ITGB1 |
| CM 4 | Fibro 6 | FN1 | ITGB1 | FN1_ITGB1 |
| CM 4 | Fibro 6 | LAMA2 | ITGB1 | LAMA2_ITGB1 |
| CM 4 | Fibro 7 | MFGE8 | PDGFRB | MFGE8_PDGFRB |
| CM 4 | Fibro 7 | TIMP1 | CD63 | TIMP1_CD63 |
| CM 4 | Fibro 7 | COL1A2 | ITGB1 | COL1A2_ITGB1 |
| CM 4 | Fibro 7 | COL3A1 | ITGB1 | COL3A1_ITGB1 |
| CM 4 | Fibro 7 | FN1 | ITGB1 | FN1_ITGB1 |
| CM 4 | Fibro 7 | LAMA2 | ITGB1 | LAMA2_ITGB1 |
| CM 4 | CM 13 | TIMP1 | CD63 | TIMP1_CD63 |
| CM 4 | CM 13 | COL1A2 | CD36 | COL1A2_CD36 |
| CM 4 | CM 13 | CALM2 | INSR | CALM2_INSR |
| CM 5 | Fibro 1 | MFGE8 | PDGFRB | MFGE8_PDGFRB |
| CM 5 | Fibro 1 | TIMP1 | CD63 | TIMP1_CD63 |
| CM 5 | Fibro 1 | LPL | LRP1 | LPL_LRP1 |
| CM 5 | Fibro 1 | PSAP | LRP1 | PSAP_LRP1 |
| CM 5 | Fibro 1 | SERPING1 | LRP1 | SERPING1_LRP1 |
| CM 5 | Fibro 7 | MFGE8 | PDGFRB | MFGE8_PDGFRB |
| CM 5 | Fibro 7 | TIMP1 | CD63 | TIMP1_CD63 |
| CM 5 | Fibro 7 | LPL | LRP1 | LPL_LRP1 |
| CM 5 | Fibro 7 | PSAP | LRP1 | PSAP_LRP1 |
| CM 5 | Fibro 7 | SERPING1 | LRP1 | SERPING1_LRP1 |
| CM 5 | Fibro 7 | COL1A1 | ITGB1 | COL1A1_ITGB1 |
| CM 5 | Fibro 7 | COL1A2 | ITGB1 | COL1A2_ITGB1 |
| CM 5 | Fibro 7 | COL3A1 | ITGB1 | COL3A1_ITGB1 |
| CM 5 | Fibro 7 | COL6A1 | ITGB1 | COL6A1_ITGB1 |
| CM 5 | Fibro 7 | COL6A2 | ITGB1 | COL6A2_ITGB1 |
| CM 5 | Fibro 7 | FN1 | ITGB1 | FN1_ITGB1 |
| CM 5 | Fibro 7 | LAMA2 | ITGB1 | LAMA2_ITGB1 |
| CM 5 | Fibro 7 | VEGFA | ITGB1 | VEGFA_ITGB1 |
| CM 7 | CM 1 | COL1A1 | CD36 | COL1A1_CD36 |
| CM 7 | CM 1 | COL1A2 | CD36 | COL1A2_CD36 |
| CM 7 | CM 1 | TIMP1 | CD63 | TIMP1_CD63 |
| CM 7 | CM 2 | COL1A1 | CD36 | COL1A1_CD36 |
| CM 7 | CM 2 | COL1A2 | CD36 | COL1A2_CD36 |
| CM 7 | CM 2 | TIMP1 | CD63 | TIMP1_CD63 |
| CM 7 | CM 2 | CALM2 | INSR | CALM2_INSR |
| CM 7 | CM 6 | COL1A1 | CD36 | COL1A1_CD36 |
| CM 7 | CM 6 | COL1A2 | CD36 | COL1A2_CD36 |
| CM 7 | CM 6 | TIMP1 | CD63 | TIMP1_CD63 |
| CM 7 | CM 6 | COL1A1 | ITGB1 | COL1A1_ITGB1 |
| CM 7 | CM 6 | COL1A2 | ITGB1 | COL1A2_ITGB1 |
| CM 7 | CM 6 | COL3A1 | ITGB1 | COL3A1_ITGB1 |
| CM 7 | CM 6 | COL6A1 | ITGB1 | COL6A1_ITGB1 |
| CM 7 | CM 6 | COL6A2 | ITGB1 | COL6A2_ITGB1 |
| CM 7 | CM 6 | FN1 | ITGB1 | FN1_ITGB1 |
| CM 7 | CM 6 | LAMA2 | ITGB1 | LAMA2_ITGB1 |
| CM 7 | CM 6 | LAMA2 | RPSA | LAMA2_RPSA |
| CM 7 | CM 9 | CALM2 | CACNA1C | CALM2_CACNA1C |
| CM 7 | CM 9 | COL1A1 | CD36 | COL1A1_CD36 |
| CM 7 | CM 9 | COL1A2 | CD36 | COL1A2_CD36 |
| CM 7 | CM 9 | TIMP1 | CD63 | TIMP1_CD63 |
| CM 7 | CM 9 | CALM2 | INSR | CALM2_INSR |
| CM 7 | CM 9 | COL1A1 | ITGB1 | COL1A1_ITGB1 |
| CM 7 | CM 9 | COL1A2 | ITGB1 | COL1A2_ITGB1 |
| CM 7 | CM 9 | COL3A1 | ITGB1 | COL3A1_ITGB1 |
| CM 7 | CM 9 | COL6A1 | ITGB1 | COL6A1_ITGB1 |
| CM 7 | CM 9 | COL6A2 | ITGB1 | COL6A2_ITGB1 |
| CM 7 | CM 9 | FN1 | ITGB1 | FN1_ITGB1 |
| CM 7 | CM 9 | LAMA2 | ITGB1 | LAMA2_ITGB1 |
| CM 7 | CM 10 | COL1A1 | CD36 | COL1A1_CD36 |
| CM 7 | CM 10 | COL1A2 | CD36 | COL1A2_CD36 |
| CM 7 | CM 10 | TIMP1 | CD63 | TIMP1_CD63 |
| CM 7 | CM 10 | COL1A1 | ITGB1 | COL1A1_ITGB1 |
| CM 7 | CM 10 | COL1A2 | ITGB1 | COL1A2_ITGB1 |
| CM 7 | CM 10 | COL3A1 | ITGB1 | COL3A1_ITGB1 |
| CM 7 | CM 10 | COL6A1 | ITGB1 | COL6A1_ITGB1 |
| CM 7 | CM 10 | COL6A2 | ITGB1 | COL6A2_ITGB1 |
| CM 7 | CM 10 | FN1 | ITGB1 | FN1_ITGB1 |
| CM 7 | CM 10 | LAMA2 | ITGB1 | LAMA2_ITGB1 |
| CM 7 | CM 10 | LAMA2 | RPSA | LAMA2_RPSA |
| CM 7 | Fibro 1 | TIMP1 | CD63 | TIMP1_CD63 |
| CM 7 | Fibro 1 | MFGE8 | PDGFRB | MFGE8_PDGFRB |
| CM 7 | Fibro 1 | PSAP | LRP1 | PSAP_LRP1 |
| CM 7 | Fibro 3 | TIMP1 | CD63 | TIMP1_CD63 |
| CM 7 | Fibro 3 | MFGE8 | PDGFRB | MFGE8_PDGFRB |
| CM 7 | Fibro 3 | PSAP | LRP1 | PSAP_LRP1 |
| CM 7 | Fibro 3 | COL1A1 | ITGB1 | COL1A1_ITGB1 |
| CM 7 | Fibro 3 | COL1A2 | ITGB1 | COL1A2_ITGB1 |
| CM 7 | Fibro 3 | COL3A1 | ITGB1 | COL3A1_ITGB1 |
| CM 7 | Fibro 3 | COL6A1 | ITGB1 | COL6A1_ITGB1 |
| CM 7 | Fibro 3 | COL6A2 | ITGB1 | COL6A2_ITGB1 |
| CM 7 | Fibro 3 | FN1 | ITGB1 | FN1_ITGB1 |
| CM 7 | Fibro 3 | LAMA2 | ITGB1 | LAMA2_ITGB1 |
| CM 7 | Fibro 4 | TIMP1 | CD63 | TIMP1_CD63 |
| CM 7 | Fibro 4 | MFGE8 | PDGFRB | MFGE8_PDGFRB |
| CM 7 | Fibro 4 | PSAP | LRP1 | PSAP_LRP1 |
| CM 7 | Fibro 4 | COL1A1 | ITGB1 | COL1A1_ITGB1 |
| CM 7 | Fibro 4 | COL1A2 | ITGB1 | COL1A2_ITGB1 |
| CM 7 | Fibro 4 | COL3A1 | ITGB1 | COL3A1_ITGB1 |
| CM 7 | Fibro 4 | COL6A1 | ITGB1 | COL6A1_ITGB1 |
| CM 7 | Fibro 4 | COL6A2 | ITGB1 | COL6A2_ITGB1 |
| CM 7 | Fibro 4 | FN1 | ITGB1 | FN1_ITGB1 |
| CM 7 | Fibro 4 | LAMA2 | ITGB1 | LAMA2_ITGB1 |
| CM 7 | Fibro 4 | FN1 | SDC2 | FN1_SDC2 |
| CM 7 | Fibro 4 | COL1A1 | ITGAV | COL1A1_ITGAV |
| CM 7 | Fibro 4 | COL1A2 | ITGAV | COL1A2_ITGAV |
| CM 7 | Fibro 4 | FN1 | ITGAV | FN1_ITGAV |
| CM 7 | Fibro 4 | MFGE8 | ITGAV | MFGE8_ITGAV |
| CM 7 | Fibro 6 | TIMP1 | CD63 | TIMP1_CD63 |
| CM 7 | Fibro 6 | MFGE8 | PDGFRB | MFGE8_PDGFRB |
| CM 7 | Fibro 6 | PSAP | LRP1 | PSAP_LRP1 |
| CM 7 | Fibro 6 | COL1A1 | ITGB1 | COL1A1_ITGB1 |
| CM 7 | Fibro 6 | COL1A2 | ITGB1 | COL1A2_ITGB1 |
| CM 7 | Fibro 6 | COL3A1 | ITGB1 | COL3A1_ITGB1 |
| CM 7 | Fibro 6 | COL6A1 | ITGB1 | COL6A1_ITGB1 |
| CM 7 | Fibro 6 | COL6A2 | ITGB1 | COL6A2_ITGB1 |
| CM 7 | Fibro 6 | FN1 | ITGB1 | FN1_ITGB1 |
| CM 7 | Fibro 6 | LAMA2 | ITGB1 | LAMA2_ITGB1 |
| CM 7 | Fibro 7 | TIMP1 | CD63 | TIMP1_CD63 |
| CM 7 | Fibro 7 | MFGE8 | PDGFRB | MFGE8_PDGFRB |
| CM 7 | Fibro 7 | PSAP | LRP1 | PSAP_LRP1 |
| CM 7 | Fibro 7 | COL1A1 | ITGB1 | COL1A1_ITGB1 |
| CM 7 | Fibro 7 | COL1A2 | ITGB1 | COL1A2_ITGB1 |
| CM 7 | Fibro 7 | COL3A1 | ITGB1 | COL3A1_ITGB1 |
| CM 7 | Fibro 7 | COL6A1 | ITGB1 | COL6A1_ITGB1 |
| CM 7 | Fibro 7 | COL6A2 | ITGB1 | COL6A2_ITGB1 |
| CM 7 | Fibro 7 | FN1 | ITGB1 | FN1_ITGB1 |
| CM 7 | Fibro 7 | LAMA2 | ITGB1 | LAMA2_ITGB1 |
| CM 7 | CM 13 | TIMP1 | CD63 | TIMP1_CD63 |
| CM 7 | CM 13 | COL1A1 | CD36 | COL1A1_CD36 |
| CM 7 | CM 13 | COL1A2 | CD36 | COL1A2_CD36 |
| CM 7 | CM 13 | CALM2 | INSR | CALM2_INSR |
| CM 11 | CM 1 | COL1A1 | CD36 | COL1A1_CD36 |
| CM 11 | CM 1 | COL1A2 | CD36 | COL1A2_CD36 |
| CM 11 | CM 1 | TIMP1 | CD63 | TIMP1_CD63 |
| CM 11 | CM 2 | COL1A1 | CD36 | COL1A1_CD36 |
| CM 11 | CM 2 | COL1A2 | CD36 | COL1A2_CD36 |
| CM 11 | CM 2 | TIMP1 | CD63 | TIMP1_CD63 |
| CM 11 | CM 2 | CALM2 | INSR | CALM2_INSR |
| CM 11 | CM 2 | SORBS1 | INSR | SORBS1_INSR |
| CM 11 | CM 6 | COL1A1 | CD36 | COL1A1_CD36 |
| CM 11 | CM 6 | COL1A2 | CD36 | COL1A2_CD36 |
| CM 11 | CM 6 | TIMP1 | CD63 | TIMP1_CD63 |
| CM 11 | CM 6 | ADAM17 | ITGB1 | ADAM17_ITGB1 |
| CM 11 | CM 6 | COL1A1 | ITGB1 | COL1A1_ITGB1 |
| CM 11 | CM 6 | COL1A2 | ITGB1 | COL1A2_ITGB1 |
| CM 11 | CM 6 | COL3A1 | ITGB1 | COL3A1_ITGB1 |
| CM 11 | CM 6 | COL6A1 | ITGB1 | COL6A1_ITGB1 |
| CM 11 | CM 6 | COL6A2 | ITGB1 | COL6A2_ITGB1 |
| CM 11 | CM 6 | FN1 | ITGB1 | FN1_ITGB1 |
| CM 11 | CM 6 | LAMA2 | RPSA | LAMA2_RPSA |
| CM 11 | CM 9 | COL1A1 | CD36 | COL1A1_CD36 |
| CM 11 | CM 9 | COL1A2 | CD36 | COL1A2_CD36 |
| CM 11 | CM 9 | TIMP1 | CD63 | TIMP1_CD63 |
| CM 11 | CM 9 | CALM2 | INSR | CALM2_INSR |
| CM 11 | CM 9 | SORBS1 | INSR | SORBS1_INSR |
| CM 11 | CM 9 | ADAM17 | ITGB1 | ADAM17_ITGB1 |
| CM 11 | CM 9 | COL1A1 | ITGB1 | COL1A1_ITGB1 |
| CM 11 | CM 9 | COL1A2 | ITGB1 | COL1A2_ITGB1 |
| CM 11 | CM 9 | COL3A1 | ITGB1 | COL3A1_ITGB1 |
| CM 11 | CM 9 | COL6A1 | ITGB1 | COL6A1_ITGB1 |
| CM 11 | CM 9 | COL6A2 | ITGB1 | COL6A2_ITGB1 |
| CM 11 | CM 9 | FN1 | ITGB1 | FN1_ITGB1 |
| CM 11 | CM 9 | LAMA2 | ITGB1 | LAMA2_ITGB1 |
| CM 11 | CM 9 | CALM2 | CACNA1C | CALM2_CACNA1C |
| CM 11 | CM 10 | COL1A1 | CD36 | COL1A1_CD36 |
| CM 11 | CM 10 | COL1A2 | CD36 | COL1A2_CD36 |
| CM 11 | CM 10 | TIMP1 | CD63 | TIMP1_CD63 |
| CM 11 | CM 10 | ADAM17 | ITGB1 | ADAM17_ITGB1 |
| CM 11 | CM 10 | COL1A1 | ITGB1 | COL1A1_ITGB1 |
| CM 11 | CM 10 | COL1A2 | ITGB1 | COL1A2_ITGB1 |
| CM 11 | CM 10 | COL3A1 | ITGB1 | COL3A1_ITGB1 |
| CM 11 | CM 10 | COL6A1 | ITGB1 | COL6A1_ITGB1 |
| CM 11 | CM 10 | COL6A2 | ITGB1 | COL6A2_ITGB1 |
| CM 11 | CM 10 | FN1 | ITGB1 | FN1_ITGB1 |
| CM 11 | CM 10 | LAMA2 | RPSA | LAMA2_RPSA |
| CM 11 | Fibro 1 | MFGE8 | PDGFRB | MFGE8_PDGFRB |
| CM 11 | Fibro 1 | LPL | LRP1 | LPL_LRP1 |
| CM 11 | Fibro 1 | PSAP | LRP1 | PSAP_LRP1 |
| CM 11 | Fibro 1 | SERPING1 | LRP1 | SERPING1_LRP1 |
| CM 11 | Fibro 1 | TIMP1 | CD63 | TIMP1_CD63 |
| CM 11 | Fibro 3 | MFGE8 | PDGFRB | MFGE8_PDGFRB |
| CM 11 | Fibro 3 | LPL | LRP1 | LPL_LRP1 |
| CM 11 | Fibro 3 | PSAP | LRP1 | PSAP_LRP1 |
| CM 11 | Fibro 3 | SERPING1 | LRP1 | SERPING1_LRP1 |
| CM 11 | Fibro 3 | TIMP1 | CD63 | TIMP1_CD63 |
| CM 11 | Fibro 3 | ADAM17 | ITGB1 | ADAM17_ITGB1 |
| CM 11 | Fibro 3 | COL1A1 | ITGB1 | COL1A1_ITGB1 |
| CM 11 | Fibro 3 | COL1A2 | ITGB1 | COL1A2_ITGB1 |
| CM 11 | Fibro 3 | COL3A1 | ITGB1 | COL3A1_ITGB1 |
| CM 11 | Fibro 3 | COL6A1 | ITGB1 | COL6A1_ITGB1 |
| CM 11 | Fibro 3 | COL6A2 | ITGB1 | COL6A2_ITGB1 |
| CM 11 | Fibro 3 | FN1 | ITGB1 | FN1_ITGB1 |
| CM 11 | Fibro 4 | MFGE8 | PDGFRB | MFGE8_PDGFRB |
| CM 11 | Fibro 4 | LPL | LRP1 | LPL_LRP1 |
| CM 11 | Fibro 4 | PSAP | LRP1 | PSAP_LRP1 |
| CM 11 | Fibro 4 | SERPING1 | LRP1 | SERPING1_LRP1 |
| CM 11 | Fibro 4 | TIMP1 | CD63 | TIMP1_CD63 |
| CM 11 | Fibro 4 | ADAM17 | ITGB1 | ADAM17_ITGB1 |
| CM 11 | Fibro 4 | COL1A1 | ITGB1 | COL1A1_ITGB1 |
| CM 11 | Fibro 4 | COL1A2 | ITGB1 | COL1A2_ITGB1 |
| CM 11 | Fibro 4 | COL3A1 | ITGB1 | COL3A1_ITGB1 |
| CM 11 | Fibro 4 | COL6A1 | ITGB1 | COL6A1_ITGB1 |
| CM 11 | Fibro 4 | COL6A2 | ITGB1 | COL6A2_ITGB1 |
| CM 11 | Fibro 4 | FN1 | ITGB1 | FN1_ITGB1 |
| CM 11 | Fibro 4 | COL1A1 | ITGAV | COL1A1_ITGAV |
| CM 11 | Fibro 4 | COL1A2 | ITGAV | COL1A2_ITGAV |
| CM 11 | Fibro 4 | FN1 | ITGAV | FN1_ITGAV |
| CM 11 | Fibro 4 | MFGE8 | ITGAV | MFGE8_ITGAV |
| CM 11 | Fibro 4 | FN1 | SDC2 | FN1_SDC2 |
| CM 11 | Fibro 6 | MFGE8 | PDGFRB | MFGE8_PDGFRB |
| CM 11 | Fibro 6 | LPL | LRP1 | LPL_LRP1 |
| CM 11 | Fibro 6 | PSAP | LRP1 | PSAP_LRP1 |
| CM 11 | Fibro 6 | SERPING1 | LRP1 | SERPING1_LRP1 |
| CM 11 | Fibro 6 | TIMP1 | CD63 | TIMP1_CD63 |
| CM 11 | Fibro 6 | ADAM17 | ITGB1 | ADAM17_ITGB1 |
| CM 11 | Fibro 6 | COL1A1 | ITGB1 | COL1A1_ITGB1 |
| CM 11 | Fibro 6 | COL1A2 | ITGB1 | COL1A2_ITGB1 |
| CM 11 | Fibro 6 | COL3A1 | ITGB1 | COL3A1_ITGB1 |
| CM 11 | Fibro 6 | COL6A1 | ITGB1 | COL6A1_ITGB1 |
| CM 11 | Fibro 6 | COL6A2 | ITGB1 | COL6A2_ITGB1 |
| CM 11 | Fibro 6 | FN1 | ITGB1 | FN1_ITGB1 |
| CM 11 | Fibro 7 | MFGE8 | PDGFRB | MFGE8_PDGFRB |
| CM 11 | Fibro 7 | LPL | LRP1 | LPL_LRP1 |
| CM 11 | Fibro 7 | PSAP | LRP1 | PSAP_LRP1 |
| CM 11 | Fibro 7 | SERPING1 | LRP1 | SERPING1_LRP1 |
| CM 11 | Fibro 7 | TIMP1 | CD63 | TIMP1_CD63 |
| CM 11 | Fibro 7 | ADAM17 | ITGB1 | ADAM17_ITGB1 |
| CM 11 | Fibro 7 | COL1A1 | ITGB1 | COL1A1_ITGB1 |
| CM 11 | Fibro 7 | COL1A2 | ITGB1 | COL1A2_ITGB1 |
| CM 11 | Fibro 7 | COL3A1 | ITGB1 | COL3A1_ITGB1 |
| CM 11 | Fibro 7 | COL6A1 | ITGB1 | COL6A1_ITGB1 |
| CM 11 | Fibro 7 | COL6A2 | ITGB1 | COL6A2_ITGB1 |
| CM 11 | Fibro 7 | FN1 | ITGB1 | FN1_ITGB1 |
| CM 11 | CM 13 | COL1A1 | CD36 | COL1A1_CD36 |
| CM 11 | CM 13 | COL1A2 | CD36 | COL1A2_CD36 |
| CM 11 | CM 13 | TIMP1 | CD63 | TIMP1_CD63 |
| CM 11 | CM 13 | CALM2 | INSR | CALM2_INSR |
| CM 11 | CM 13 | SORBS1 | INSR | SORBS1_INSR |

### Supplemental Table ST6. Ligands Lost in HCM Fibroblast Cluster 4 and Associated Molecular Functions

| | **Ligands lost in HCM fibro 4 condition** | | | | | | | | | | | | | | | | | | | | | | | | |
|---|---|---|---|---|---|---|---|---|---|---|---|---|---|---|---|---|---|---|---|---|---|---|---|---|---|
| **Molecular Function** | C3 | CALR | COL18A1 | COL4A1 | COL5A1 | COL5A2 | CXCL12 | CYR61 | FBLN1 | FBN1 | HSP90B1 | IGF1 | LAMA2 | LAMB1 | LAMC1 | LGALS3BP | LRPAP1 | NID1 | PDGFD | PTN | TFPI | THBS2 | TIMP2 | TNC | **Count** |
| amide binding | | x | | | | | | | | | | | | | | | x | | | | | | | | 2 |
| collagen binding | | | | | | | | | | | | | | | | | | x | | | | | | | 1 |
| endopeptidase inhibitor activity | x | | | | | | | | | | | | | | | | | | | | x | | x | | 3 |
| endopeptidase regulator activity | x | | | | | | | | | | | | | | | | | | | | x | | x | | 3 |
| enzyme inhibitor activity | x | | | | | | | | | | | | | | | | | | | x | x | | x | | 4 |
| extracellular matrix binding | | | | | | | | | | | | | | | | | | x | | | | | | | 1 |
| extracellular matrix structural constituent | | | x | x | x | x | | | x | | | | | x | | | | x | | | | | | | 7 |
| extracellular matrix structural constituent conferring tensile strength | | | x | x | x | x | | | | | | | x | | | | | | | | | | | | 5 |
| glycosaminoglycan binding | | | | | x | | | | | | | | | | | | x | | | x | | | | | 3 |
| growth factor activity | | | | | | | | | | | | x | | | | | | | | x | | | | | 2 |
| growth factor binding | | | | x | x | x | | | | | | | | | | | | | | x | | | | | 4 |
| heparin binding | | | | | x | | | | | | | | | | | | x | | | x | | | | | 3 |
| insulin-like growth factor receptor binding | | | | | | | | | | | | x | | | | | | | | | | | | | 1 |
| integrin binding | | x | | | x | | | | | | | x | | x | | | | | | x | | | x | | 6 |
| lipoprotein particle receptor binding | | | | | | | | | | | x | | | | | | x | | | | | | | | 2 |
| low-density lipoprotein particle receptor binding | | | | | | | | | | | x | | | | | | x | | | | | | | | 2 |
| metalloendopeptidase inhibitor activity | | | | | | | | | | | | | | | | | | | | | | | x | | 1 |
| peptidase activator activity | | | | | | | | | x | | | | | | | | | | | | | | | | 1 |
| peptidase inhibitor activity | x | | | | | | | | | | | | | | | | | | | | x | | x | | 3 |
| peptidase regulator activity | x | | | | | | | | x | | | | | | | | | | | | x | | x | | 4 |
| peptide binding | | x | | | | | | | | | | | | | | | x | | | | | | | | 2 |
| phosphatase regulator activity | | | | | | | | | | | | | | | | | | | | x | | | | | 1 |
| platelet-derived growth factor binding | | | | x | x | | | | | | | | | | | | | | | | | | | | 2 |
| protease binding | | | | | | | | | | | | | | | | | | | | | | | x | | 1 |
| protein C-terminus binding | | | | | | | | | x | | | | | | | | | | | | | | | | 1 |
| protein phosphatase regulator activity | | | | | | | | | | | | | | | | | | | | x | | | | | 1 |
| proteoglycan binding | | | | | x | | | | | | | | | | | | | x | | x | | | | | 3 |
| receptor ligand activity | | | | | | | | | | | | x | | | | | x | | | x | | | | | 3 |
| signaling receptor activator activity | | | | | | | | | | | | x | | | | | x | | | x | | | | | 3 |
| SMAD binding | | | | | | x | | | | | | | | | | | | | | | | | | | 1 |
| sulfur compound binding | | | | | x | | | | | | | | | | | | x | | | x | | | | | 3 |

### Supplemental Table ST7. Reduction in Fibroblast Cluster 4 Ligand-Receptor Interactions in HCM

| Ligand | Receptor | L-R pairs | HCM |
|---|---|---|---|
| Fibro 4 | Fibro 3 | 41 | 10 |
| Fibro 2 | Fibro 4 | 51 | 18 |
| Fibro 4 | Fibro 8 | 39 | 4 |
| Fibro 4 | Fibro 7 | 39 | 3 |
| Fibro 4 | Fibro 4 | 54 | 12 |

### Supplemental Table ST8. Ligand-Receptor Interactions that are Increased in HCM between Fibroblast Subtypes and Cardiomyocyte Subtypes

Normal

| L-Cell | R-Cell | L | R | L-R pair |
|---|---|---|---|---|
| Fibro 2 | CM 1 | C3 | CD81 | C3_CD81 |
| Fibro 2 | CM 1 | TIMP1 | CD63 | TIMP1_CD63 |
| Fibro 2 | CM 1 | COL1A1 | CD36 | COL1A1_CD36 |
| Fibro 2 | CM 1 | COL1A2 | CD36 | COL1A2_CD36 |
| Fibro 3 | CM 1 | C3 | CD81 | C3_CD81 |
| Fibro 3 | CM 1 | TIMP1 | CD63 | TIMP1_CD63 |
| Fibro 3 | CM 1 | COL1A1 | CD36 | COL1A1_CD36 |
| Fibro 3 | CM 1 | COL1A2 | CD36 | COL1A2_CD36 |
| Fibro 4 | CM 1 | C3 | CD81 | C3_CD81 |
| Fibro 4 | CM 1 | TIMP1 | CD63 | TIMP1_CD63 |
| Fibro 4 | CM 1 | COL1A1 | CD36 | COL1A1_CD36 |
| Fibro 4 | CM 1 | COL1A2 | CD36 | COL1A2_CD36 |
| Fibro 5 | CM 1 | C3 | CD81 | C3_CD81 |
| Fibro 5 | CM 1 | TIMP1 | CD63 | TIMP1_CD63 |
| Fibro 5 | CM 1 | COL1A1 | CD36 | COL1A1_CD36 |
| Fibro 5 | CM 1 | COL1A2 | CD36 | COL1A2_CD36 |
| Fibro 2 | CM 2 | TIMP1 | CD63 | TIMP1_CD63 |
| Fibro 2 | CM 2 | COL1A1 | CD36 | COL1A1_CD36 |
| Fibro 2 | CM 2 | COL1A2 | CD36 | COL1A2_CD36 |
| Fibro 2 | CM 2 | CALM2 | INSR | CALM2_INSR |
| Fibro 2 | CM 2 | IGF1 | INSR | IGF1_INSR |
| Fibro 3 | CM 2 | TIMP1 | CD63 | TIMP1_CD63 |
| Fibro 3 | CM 2 | COL1A1 | CD36 | COL1A1_CD36 |
| Fibro 3 | CM 2 | COL1A2 | CD36 | COL1A2_CD36 |
| Fibro 3 | CM 2 | CALM2 | INSR | CALM2_INSR |
| Fibro 3 | CM 2 | IGF1 | INSR | IGF1_INSR |
| Fibro 4 | CM 2 | TIMP1 | CD63 | TIMP1_CD63 |
| Fibro 4 | CM 2 | COL1A1 | CD36 | COL1A1_CD36 |
| Fibro 4 | CM 2 | COL1A2 | CD36 | COL1A2_CD36 |
| Fibro 4 | CM 2 | CALM2 | INSR | CALM2_INSR |
| Fibro 4 | CM 2 | IGF1 | INSR | IGF1_INSR |
| Fibro 5 | CM 2 | TIMP1 | CD63 | TIMP1_CD63 |
| Fibro 5 | CM 2 | COL1A1 | CD36 | COL1A1_CD36 |
| Fibro 5 | CM 2 | COL1A2 | CD36 | COL1A2_CD36 |
| Fibro 5 | CM 2 | CALM2 | INSR | CALM2_INSR |
| Fibro 5 | CM 2 | IGF1 | INSR | IGF1_INSR |

HCM

| L-Cell | R-Cell | L | R | L-R pair |
|---|---|---|---|---|
| Fibro 2 | CM 1 | COL1A1 | CD36 | COL1A1_CD36 |
| Fibro 2 | CM 1 | COL1A2 | CD36 | COL1A2_CD36 |
| Fibro 2 | CM 1 | COL1A1 | ITGB1 | COL1A1_ITGB1 |
| Fibro 2 | CM 1 | COL1A2 | ITGB1 | COL1A2_ITGB1 |
| Fibro 2 | CM 1 | COL3A1 | ITGB1 | COL3A1_ITGB1 |
| Fibro 2 | CM 1 | COL6A1 | ITGB1 | COL6A1_ITGB1 |
| Fibro 2 | CM 1 | COL6A2 | ITGB1 | COL6A2_ITGB1 |
| Fibro 2 | CM 1 | COL6A3 | ITGB1 | COL6A3_ITGB1 |
| Fibro 2 | CM 1 | FBLN1 | ITGB1 | FBLN1_ITGB1 |
| Fibro 2 | CM 1 | FBN1 | ITGB1 | FBN1_ITGB1 |
| Fibro 2 | CM 1 | FN1 | ITGB1 | FN1_ITGB1 |
| Fibro 2 | CM 1 | HSPG2 | ITGB1 | HSPG2_ITGB1 |
| Fibro 2 | CM 1 | LAMA2 | ITGB1 | LAMA2_ITGB1 |
| Fibro 2 | CM 1 | LAMC1 | ITGB1 | LAMC1_ITGB1 |
| Fibro 2 | CM 1 | LGALS3BP | ITGB1 | LGALS3BP_ITGB1 |
| Fibro 2 | CM 1 | TIMP2 | ITGB1 | TIMP2_ITGB1 |
| Fibro 2 | CM 1 | VCAN | ITGB1 | VCAN_ITGB1 |
| Fibro 2 | CM 1 | TIMP1 | CD63 | TIMP1_CD63 |
| Fibro 3 | CM 1 | COL1A2 | CD36 | COL1A2_CD36 |
| Fibro 3 | CM 1 | COL1A2 | ITGB1 | COL1A2_ITGB1 |
| Fibro 3 | CM 1 | COL3A1 | ITGB1 | COL3A1_ITGB1 |
| Fibro 3 | CM 1 | COL6A1 | ITGB1 | COL6A1_ITGB1 |
| Fibro 3 | CM 1 | COL6A2 | ITGB1 | COL6A2_ITGB1 |
| Fibro 3 | CM 1 | COL6A3 | ITGB1 | COL6A3_ITGB1 |
| Fibro 3 | CM 1 | FBLN1 | ITGB1 | FBLN1_ITGB1 |
| Fibro 3 | CM 1 | FN1 | ITGB1 | FN1_ITGB1 |
| Fibro 3 | CM 1 | HSPG2 | ITGB1 | HSPG2_ITGB1 |
| Fibro 3 | CM 1 | LAMC1 | ITGB1 | LAMC1_ITGB1 |
| Fibro 3 | CM 1 | VCAN | ITGB1 | VCAN_ITGB1 |
| Fibro 3 | CM 1 | TIMP1 | CD63 | TIMP1_CD63 |
| Fibro 4 | CM 1 | COL1A1 | CD36 | COL1A1_CD36 |
| Fibro 4 | CM 1 | COL1A2 | CD36 | COL1A2_CD36 |
| Fibro 4 | CM 1 | COL1A1 | ITGB1 | COL1A1_ITGB1 |
| Fibro 4 | CM 1 | COL1A2 | ITGB1 | COL1A2_ITGB1 |
| Fibro 4 | CM 1 | COL3A1 | ITGB1 | COL3A1_ITGB1 |
| Fibro 4 | CM 1 | COL6A1 | ITGB1 | COL6A1_ITGB1 |
| Fibro 4 | CM 1 | COL6A2 | ITGB1 | COL6A2_ITGB1 |
| Fibro 4 | CM 1 | COL6A3 | ITGB1 | COL6A3_ITGB1 |
| Fibro 4 | CM 1 | FN1 | ITGB1 | FN1_ITGB1 |
| Fibro 4 | CM 1 | HSPG2 | ITGB1 | HSPG2_ITGB1 |
| Fibro 4 | CM 1 | VCAN | ITGB1 | VCAN_ITGB1 |
| Fibro 4 | CM 1 | TIMP1 | CD63 | TIMP1_CD63 |
| Fibro 5 | CM 1 | COL1A1 | CD36 | COL1A1_CD36 |
| Fibro 5 | CM 1 | COL1A2 | CD36 | COL1A2_CD36 |
| Fibro 5 | CM 1 | COL1A1 | ITGB1 | COL1A1_ITGB1 |
| Fibro 5 | CM 1 | COL1A2 | ITGB1 | COL1A2_ITGB1 |
| Fibro 5 | CM 1 | COL3A1 | ITGB1 | COL3A1_ITGB1 |
| Fibro 5 | CM 1 | COL6A1 | ITGB1 | COL6A1_ITGB1 |
| Fibro 5 | CM 1 | COL6A2 | ITGB1 | COL6A2_ITGB1 |
| Fibro 5 | CM 1 | COL6A3 | ITGB1 | COL6A3_ITGB1 |
| Fibro 5 | CM 1 | FN1 | ITGB1 | FN1_ITGB1 |
| Fibro 5 | CM 1 | HSPG2 | ITGB1 | HSPG2_ITGB1 |
| Fibro 5 | CM 1 | LAMA2 | ITGB1 | LAMA2_ITGB1 |
| Fibro 5 | CM 1 | LAMB1 | ITGB1 | LAMB1_ITGB1 |
| Fibro 5 | CM 1 | LAMC1 | ITGB1 | LAMC1_ITGB1 |
| Fibro 5 | CM 1 | VCAN | ITGB1 | VCAN_ITGB1 |
| Fibro 5 | CM 1 | TIMP1 | CD63 | TIMP1_CD63 |
| Fibro 2 | CM 2 | COL1A1 | CD36 | COL1A1_CD36 |
| Fibro 2 | CM 2 | COL1A2 | CD36 | COL1A2_CD36 |
| Fibro 2 | CM 2 | COL1A1 | ITGB1 | COL1A1_ITGB1 |
| Fibro 2 | CM 2 | COL1A2 | ITGB1 | COL1A2_ITGB1 |
| Fibro 2 | CM 2 | COL3A1 | ITGB1 | COL3A1_ITGB1 |
| Fibro 2 | CM 2 | COL6A1 | ITGB1 | COL6A1_ITGB1 |
| Fibro 2 | CM 2 | COL6A2 | ITGB1 | COL6A2_ITGB1 |
| Fibro 2 | CM 2 | COL6A3 | ITGB1 | COL6A3_ITGB1 |
| Fibro 2 | CM 2 | FBLN1 | ITGB1 | FBLN1_ITGB1 |
| Fibro 2 | CM 2 | FBN1 | ITGB1 | FBN1_ITGB1 |
| Fibro 2 | CM 2 | FN1 | ITGB1 | FN1_ITGB1 |
| Fibro 2 | CM 2 | HSPG2 | ITGB1 | HSPG2_ITGB1 |
| Fibro 2 | CM 2 | LAMA2 | ITGB1 | LAMA2_ITGB1 |
| Fibro 2 | CM 2 | LAMC1 | ITGB1 | LAMC1_ITGB1 |
| Fibro 2 | CM 2 | LGALS3BP | ITGB1 | LGALS3BP_ITGB1 |
| Fibro 2 | CM 2 | TIMP2 | ITGB1 | TIMP2_ITGB1 |
| Fibro 2 | CM 2 | VCAN | ITGB1 | VCAN_ITGB1 |
| Fibro 2 | CM 2 | TIMP1 | CD63 | TIMP1_CD63 |
| Fibro 3 | CM 2 | COL1A2 | CD36 | COL1A2_CD36 |
| Fibro 3 | CM 2 | COL1A2 | ITGB1 | COL1A2_ITGB1 |
| Fibro 3 | CM 2 | COL3A1 | ITGB1 | COL3A1_ITGB1 |
| Fibro 3 | CM 2 | COL6A1 | ITGB1 | COL6A1_ITGB1 |
| Fibro 3 | CM 2 | COL6A2 | ITGB1 | COL6A2_ITGB1 |
| Fibro 3 | CM 2 | COL6A3 | ITGB1 | COL6A3_ITGB1 |
| Fibro 3 | CM 2 | FBLN1 | ITGB1 | FBLN1_ITGB1 |
| Fibro 3 | CM 2 | FN1 | ITGB1 | FN1_ITGB1 |
| Fibro 3 | CM 2 | HSPG2 | ITGB1 | HSPG2_ITGB1 |
| Fibro 3 | CM 2 | LAMC1 | ITGB1 | LAMC1_ITGB1 |
| Fibro 3 | CM 2 | VCAN | ITGB1 | VCAN_ITGB1 |
| Fibro 3 | CM 2 | TIMP1 | CD63 | TIMP1_CD63 |
| Fibro 4 | CM 2 | COL1A1 | CD36 | COL1A1_CD36 |
| Fibro 4 | CM 2 | COL1A2 | CD36 | COL1A2_CD36 |
| Fibro 4 | CM 2 | COL1A1 | ITGB1 | COL1A1_ITGB1 |
| Fibro 4 | CM 2 | COL1A2 | ITGB1 | COL1A2_ITGB1 |
| Fibro 4 | CM 2 | COL3A1 | ITGB1 | COL3A1_ITGB1 |
| Fibro 4 | CM 2 | COL6A1 | ITGB1 | COL6A1_ITGB1 |
| Fibro 4 | CM 2 | COL6A2 | ITGB1 | COL6A2_ITGB1 |
| Fibro 4 | CM 2 | COL6A3 | ITGB1 | COL6A3_ITGB1 |
| Fibro 4 | CM 2 | FN1 | ITGB1 | FN1_ITGB1 |
| Fibro 4 | CM 2 | HSPG2 | ITGB1 | HSPG2_ITGB1 |
| Fibro 4 | CM 2 | VCAN | ITGB1 | VCAN_ITGB1 |
| Fibro 4 | CM 2 | TIMP1 | CD63 | TIMP1_CD63 |
| Fibro 5 | CM 2 | COL1A1 | CD36 | COL1A1_CD36 |
| Fibro 5 | CM 2 | COL1A2 | CD36 | COL1A2_CD36 |
| Fibro 5 | CM 2 | COL1A1 | ITGB1 | COL1A1_ITGB1 |
| Fibro 5 | CM 2 | COL1A2 | ITGB1 | COL1A2_ITGB1 |
| Fibro 5 | CM 2 | COL3A1 | ITGB1 | COL3A1_ITGB1 |
| Fibro 5 | CM 2 | COL6A1 | ITGB1 | COL6A1_ITGB1 |
| Fibro 5 | CM 2 | COL6A2 | ITGB1 | COL6A2_ITGB1 |
| Fibro 5 | CM 2 | COL6A3 | ITGB1 | COL6A3_ITGB1 |
| Fibro 5 | CM 2 | FN1 | ITGB1 | FN1_ITGB1 |
| Fibro 5 | CM 2 | HSPG2 | ITGB1 | HSPG2_ITGB1 |
| Fibro 5 | CM 2 | LAMA2 | ITGB1 | LAMA2_ITGB1 |
| Fibro 5 | CM 2 | LAMB1 | ITGB1 | LAMB1_ITGB1 |
| Fibro 5 | CM 2 | LAMC1 | ITGB1 | LAMC1_ITGB1 |
| Fibro 5 | CM 2 | VCAN | ITGB1 | VCAN_ITGB1 |

### Supplemental Table ST9. Ligand-Receptor Interactions that are Increased in HCM between Cardiomyocyte Cluster 8 and Cardiomyocyte Subtypes and Myofibroblasts

Normal

| L-Cell | R-Cell | L | R | L-R pair |
|---|---|---|---|---|
| CM 8 | CM 1 | COL1A1 | CD36 | COL1A1_CD36 |
| CM 8 | CM 1 | COL1A2 | CD36 | COL1A2_CD36 |
| CM 8 | CM 1 | TIMP1 | CD63 | TIMP1_CD63 |
| CM 8 | CM 1 | C3 | CD81 | C3_CD81 |
| CM 8 | CM 2 | COL1A1 | CD36 | COL1A1_CD36 |
| CM 8 | CM 2 | COL1A2 | CD36 | COL1A2_CD36 |
| CM 8 | CM 2 | TIMP1 | CD63 | TIMP1_CD63 |
| CM 8 | CM 2 | CALM2 | INSR | CALM2_INSR |
| CM 8 | CM 2 | IGF1 | INSR | IGF1_INSR |
| CM 8 | CM 13 | COL1A2 | CD36 | COL1A2_CD36 |
| CM 8 | CM 13 | TIMP1 | CD63 | TIMP1_CD63 |
| CM 8 | CM 13 | CALM2 | INSR | CALM2_INSR |
| CM 8 | CM 13 | IGF1 | INSR | IGF1_INSR |

HCM

| L-Cell | R-Cell | L | R | L-R pair |
|---|---|---|---|---|
| CM 8 | CM 1 | COL1A2 | CD36 | COL1A2_CD36 |
| CM 8 | CM 1 | TIMP1 | CD63 | TIMP1_CD63 |
| CM 8 | CM 1 | COL1A2 | ITGB1 | COL1A2_ITGB1 |
| CM 8 | CM 1 | COL3A1 | ITGB1 | COL3A1_ITGB1 |
| CM 8 | CM 1 | COL6A1 | ITGB1 | COL6A1_ITGB1 |
| CM 8 | CM 1 | COL6A2 | ITGB1 | COL6A2_ITGB1 |
| CM 8 | CM 1 | FN1 | ITGB1 | FN1_ITGB1 |
| CM 8 | CM 1 | HSPG2 | ITGB1 | HSPG2_ITGB1 |
| CM 8 | CM 1 | LAMA2 | ITGB1 | LAMA2_ITGB1 |
| CM 8 | CM 1 | LGALS3BP | ITGB1 | LGALS3BP_ITGB1 |
| CM 8 | CM 2 | COL1A2 | CD36 | COL1A2_CD36 |
| CM 8 | CM 2 | TIMP1 | CD63 | TIMP1_CD63 |
| CM 8 | CM 2 | COL1A2 | ITGB1 | COL1A2_ITGB1 |
| CM 8 | CM 2 | COL3A1 | ITGB1 | COL3A1_ITGB1 |
| CM 8 | CM 2 | COL6A1 | ITGB1 | COL6A1_ITGB1 |
| CM 8 | CM 2 | COL6A2 | ITGB1 | COL6A2_ITGB1 |
| CM 8 | CM 2 | FN1 | ITGB1 | FN1_ITGB1 |
| CM 8 | CM 2 | HSPG2 | ITGB1 | HSPG2_ITGB1 |
| CM 8 | CM 2 | LAMA2 | ITGB1 | LAMA2_ITGB1 |
| CM 8 | CM 2 | LGALS3BP | ITGB1 | LGALS3BP_ITGB1 |
| CM 8 | CM 13 | COL1A2 | CD36 | COL1A2_CD36 |
| CM 8 | CM 13 | TIMP1 | CD63 | TIMP1_CD63 |
| CM 8 | CM 13 | COL1A2 | ITGB1 | COL1A2_ITGB1 |
| CM 8 | CM 13 | COL3A1 | ITGB1 | COL3A1_ITGB1 |
| CM 8 | CM 13 | COL6A1 | ITGB1 | COL6A1_ITGB1 |
| CM 8 | CM 13 | COL6A2 | ITGB1 | COL6A2_ITGB1 |
| CM 8 | CM 13 | FN1 | ITGB1 | FN1_ITGB1 |
| CM 8 | CM 13 | HSPG2 | ITGB1 | HSPG2_ITGB1 |
| CM 8 | CM 13 | LAMA2 | ITGB1 | LAMA2_ITGB1 |
| CM 8 | CM 13 | LGALS3BP | ITGB1 | LGALS3BP_ITGB1 |
